# One Size Does Not Fit All: A Data-Driven Framework for Personalized Comorbidity Scoring

**DOI:** 10.64898/2026.06.30.26356967

**Authors:** Yeon-Mi Hwang, Ying Cui, Javen Xu, Tianyu Pan, Ron Li, Brian T. Rice, Lu Tian, Tina Hernandez-Boussard

## Abstract

Comorbidity indices are widely used in clinical research to summarize disease burden. However, traditional indices were developed decades ago in limited populations using fixed weights that do not reflect the diversity of patients in modern healthcare. We present the Personalized Comorbidity Score (PCS), a data-driven framework for context-dependent comorbidity scoring designed to capture patient complexity while remaining accessible for broad research adoption. PCS was developed using Epic Cosmos, a large national EHR network encompassing over 8 million adult inpatient encounters from 2015 to 2020, with comorbidities defined using AHRQ Clinical Classifications Software Refined categories. Models were developed separately across eight age-sex subgroups using LASSO-penalized Cox regression for feature selection and restricted mean survival time for score derivation. PCS is available in two versions: PCS Core, incorporating age, sex, and comorbidities, and PCS Extended, which additionally incorporates socioeconomic and geographic variables. PCS Core and PCS Extended achieved AUROCs of 0.812 and 0.813 for one-year mortality, outperforming traditional indices (AUROC, 0.714-0.730). PCS demonstrated consistently lower subgroup calibration error across demographic and socioeconomic groups without including race or ethnicity as model features. PCS was further evaluated in two complementary external EHR datasets (Stanford Health Care and MIMIC-IV) with distinct patient populations and data structures, where it consistently outperformed traditional indices. Open-source R and Python packages are provided to support broad adoption. PCS provides an updatable framework for comorbidity measurement that is accurate, context-dependent, and designed to evolve alongside clinical practice.

## 1. Introduction

Comorbidity indices underpin medical research by efficiently summarizing a patient’s burden of multiple chronic conditions into a single numeric score.^1–3^ They quantify disease burden for a range of research and administrative applications, including risk adjustment, clinical prediction, resource planning, and reimbursement.^1,3,4^ Of these, the Charlson Comorbidity Index (CCI) dominates the field. The CCI was developed in 1987 from approximately 600 patient records to predict one-year mortality, and numerous adaptations have since been developed.^5–7^ The Elixhauser Comorbidity Index (ECI) is the most widely used alternative. Originally developed in 1998 as a set of 30 binary condition flags using administrative inpatient data, it was later converted into several weighted scoring systems, of which the van Walraven ECI is the most widely used.^8,9^ Although these indices were developed to predict specific outcomes in inpatient populations, they have been extensively validated across diverse cohorts and outcomes.^5,8,9^ Their simplicity, versatility, and consistent performance have made them the standard tools in medical research. However, as the diversity of patients and complexity of modern healthcare have grown, important limitations have emerged.

Four limitations have become apparent over time: uniform weights that fail to reflect population- and outcome-specific differences, incomplete condition coverage, lack of socioeconomic context, and fragmented updating. First, traditional indices apply uniform weights regardless of a patient’s age, sex, or the outcome being measured. Yet the prognostic significance of comorbid conditions varies considerably across these dimensions. Age and sex are well-established modifiers of disease burden, and comorbidity profiles differ across specific populations, which general-purpose indices are not designed to capture. Cohort-specific comorbidity indices illustrate this point.^10–12^ Beyond population differences, the same condition can carry fundamentally different prognostic significance depending on the outcome being measured. For example, diabetes with chronic complications carries a weight of -3 for mortality but 9 for readmission in the Moore ECI.^13^ As a result, traditional indices may systematically misrepresent disease burden for patients who differ by age, sex, population context, or outcomes of interest, with direct consequences for the fairness of comorbidity measurement in research.

Second, the conditions these indices capture are incomplete. Our understanding of disease burden has expanded considerably since the CCI and ECI were developed. Mental health and behavioral conditions are now well-established predictors of mortality and healthcare utilization, yet they are largely absent from the CCI, which remains the dominant index in practice.^14^ Beyond established conditions, emerging ones such as long COVID highlight the need for frameworks that can systematically incorporate new disease entities as medical knowledge evolves.^15^ Incomplete condition coverage can disproportionately affect populations whose disease burden falls outside what traditional indices capture.

Third, traditional indices do not account for the socioeconomic context in which patients live and receive care. Socioeconomic factors are well-established determinants of health outcomes.^16^ A patient with poorly controlled diabetes in a rural area with no insurance may face a vastly different disease burden than one with the same diagnosis in an urban center with full coverage, yet both receive the same comorbidity score. These contextual differences are invisible to traditional indices. This limitation is particularly consequential given well-documented health inequalities across age, sex, race, ethnicity, and socioeconomic groups. Comorbidity tools that fail to account for population heterogeneity may systematically misrepresent disease burden in already underserved populations.

Fourth, updates to traditional indices have been fragmented, leaving researchers without clear guidance on which version to use. The prognostic significance of conditions can shift substantially as treatments evolve. HIV/AIDS was assigned a weight of 6 in the original Charlson index, reflecting its severe mortality burden before effective antiretroviral therapy existed.^1,5,17^ Successive updates have consistently reduced that weight as treatment outcomes improved. Nevertheless, the original Charlson index remains one of the most cited comorbidity tools in the literature.^5^ Without a systematic updating process, version proliferation undermines reproducibility and comparability across studies.

Together, these gaps reflect a fundamental limitation. Traditional indices treat comorbidity burden as fixed and universal, when in reality it is context-dependent and population-specific. Advances in electronic health records (EHRs) and large-scale data infrastructure now make it possible to address these limitations directly. We present the Personalized Comorbidity Score (PCS) as a demonstration of a generalizable and data-driven framework for context-dependent comorbidity scoring, designed to address each of these gaps. A core design motivation was to capture individual patient complexity through context-dependent scoring, while remaining simple enough for broad research adoption. PCS was built on a nationally diverse training population with comprehensive condition coverage, age-sex stratified comorbidity selection and weighting, and explicit incorporation of socioeconomic context. Performance across demographic and socioeconomic subgroups was evaluated alongside discrimination, with subgroup calibration serving as the primary fairness evaluation metric. PCS is intended for use by researchers as a summary covariate for risk adjustment and downstream prediction models. Researchers may apply the pre-trained weights directly or use the framework to derive context-specific weights.

We hypothesized that a personalized, context-aware approach to comorbidity scoring would achieve better discrimination and greater fairness across demographic subgroups compared to existing indices. As a demonstration of this framework, we developed and evaluated PCS using Epic Cosmos, a national EHR network encompassing over 300 million patients, with comorbidities defined using AHRQ Clinical Classifications Software Refined (CCSR) categories. PCS was developed on inpatient populations for one-year mortality prediction, mirroring the original CCI design.^5^ PCS is available in two versions: PCS Core, stratified by age and sex, and PCS Extended, which additionally incorporates socioeconomic measures. To support broad adoption, open-source R and Python packages have been developed and will be made publicly available, enabling researchers to apply PCS directly from CCSR-coded diagnoses. This work represents a step toward comorbidity measurement that is more accurate, more context-aware, and designed to evolve alongside both clinical practice and medical knowledge.

## 2. Methods

An overview of the PCS framework is provided in Figure 1.

**Figure 1.**
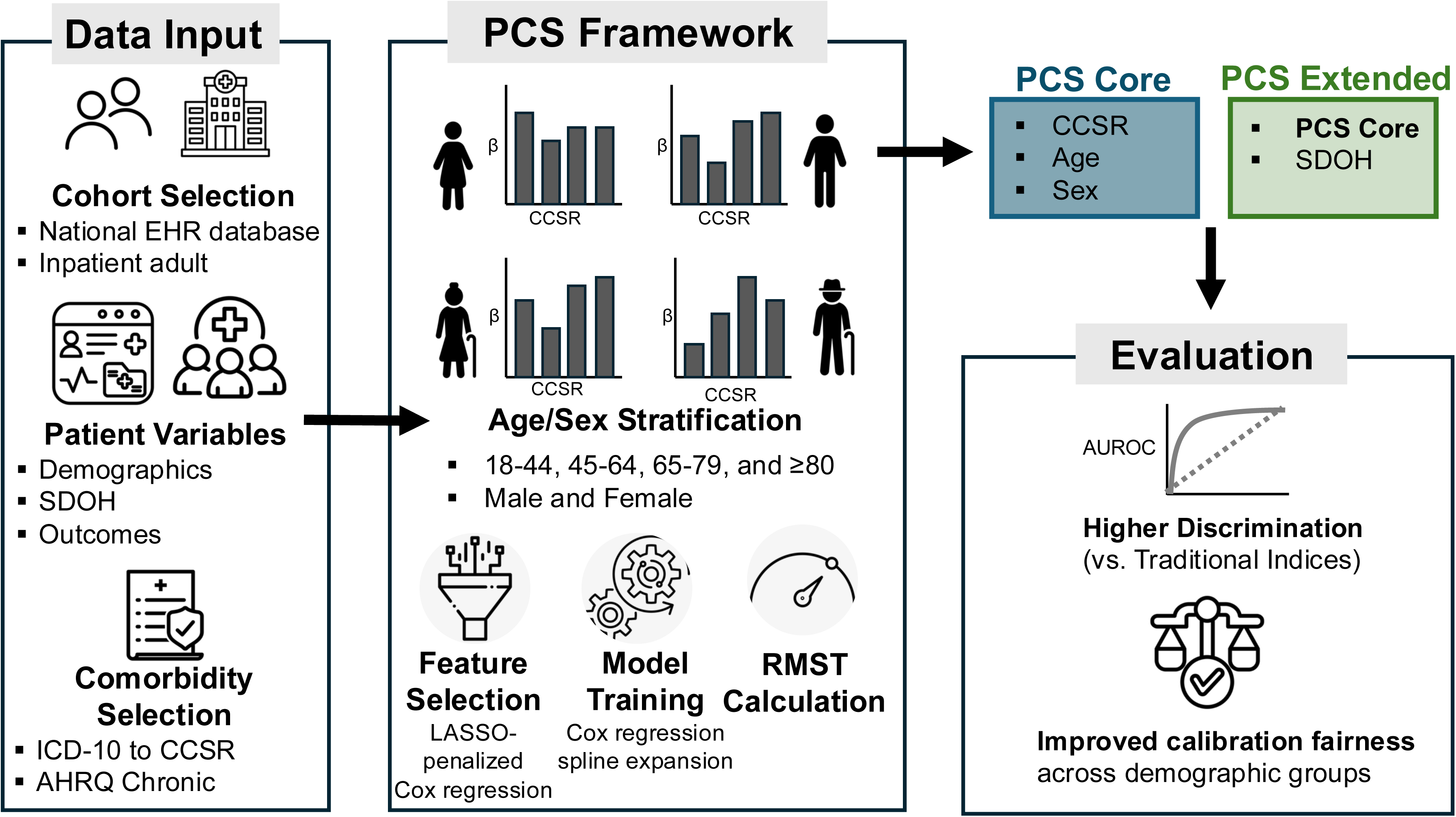
Overview of the Personalized Comorbidity Score (PCS) framework. Abbreviations: AHRQ, Agency for Healthcare Research and Quality; CCSR, Clinical Classifications Software Refined; ICD-10, International Classification of Diseases, Tenth Revision; LASSO, Least Absolute Shrinkage and Selection Operator; PCS, Personalized Comorbidity Score; SDOH, Social Determinants of Health. Comorbidities were identified from ICD-10 codes mapped to AHRQ CCSR chronic categories. Models were developed separately across eight age-sex subgroups using LASSO-penalized Cox regression for feature selection and RMST-based score derivation. PCS Core incorporates CCSR-defined comorbidities, age, and sex. PCS Extended additionally incorporates socioeconomic and geographic variables.

### 2.1 Study population

This retrospective cohort study used de-identified EHR data from Epic Cosmos, a national research database encompassing 307 million patients across 2197 hospitals and 49,300 clinics in the United States (as of June 29, 2026).^18^ Data for this study were accessed on October 13, 2025. Participating organizations collectively serve a patient population whose demographic composition is similar to U.S. Census distributions, though Epic Cosmos predominantly captures patients receiving care within the Epic network and may underrepresent uninsured individuals and those treated at non-Epic facilities.^18,19^ We identified adult patients (aged ≥18 years) with hospital admissions between March 31, 2015, and March 31, 2020. This observation period was selected to ensure sufficient EHR adoption and data quality and to allow a minimum of five years of follow-up for the assessment of long-term mortality outcomes. To ensure adequate capture of pre-existing comorbidities, we required patients to have at least two healthcare encounters prior to the index admission with a minimum care span of 365 days. We excluded admissions with zero-day length of stay, non-index admissions, and pregnancy-related admissions. For patients with multiple qualifying admissions during the study period, only the first qualifying admission was included. The full cohort selection process is described in Figure 2.

**Figure 2.**
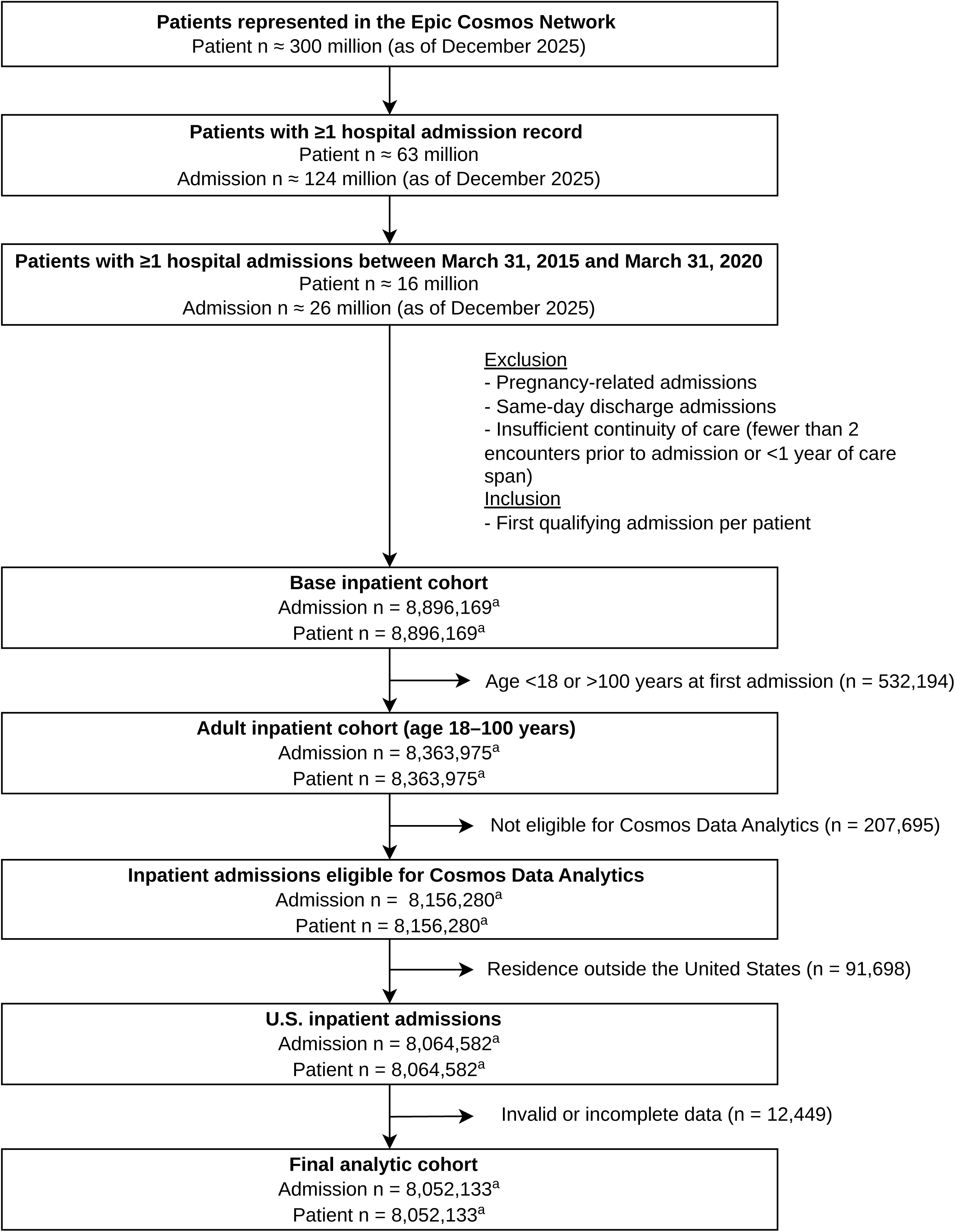
Cohort selection flowchart for the PCS development. Starting from approximately 300 million patients in the Epic Cosmos network, eligible inpatient admissions between March 31, 2015 and March 31, 2020 were identified and sequentially filtered. Exclusions were applied for pregnancy-related admissions, same-day discharges, and insufficient continuity of care. Only the first qualifying admission per patient was retained. Further exclusions were applied for age outside the 18–100 year range, ineligibility for Cosmos Data Analytics, residence outside the United States, and invalid or incomplete data, yielding a final analytic cohort of 8,052,133 admissions. ^a^Data Extract Date: October 13, 2025.

### 2.2 Variables

#### 2.2.1 Patient characteristics

We extracted patient demographic characteristics including age at admission, sex, race, ethnicity, multiracial status, and insurance type. Neighborhood-level socioeconomic status and geographic classification were assessed using the Social Vulnerability Index (SVI) and Rural-Urban Commuting Area (RUCA) code. All variables were obtained directly from Epic Cosmos without modification (Table S1).

Records with missing values in predictors required for each model specification were excluded from model development (complete-case analysis). Missingness for demographic and socioeconomic variables is reported in Table 1.

**Table 1.** Demographic and clinical characteristics of the full analytic cohort and the SneakPeek 1% held-out validation subset.

|  | Missing | Epic Cosmos<br>SneakPeek<br>(1% Dataset; n=80,816) | Missing | Epic Cosmos<br>(n=8,052,133) | SMD |
| --- | --- | --- | --- | --- | --- |
| **Patient Characteristics** |  |  |  |  |  |
| Age, mean (SD) | 0 | 63.43 ± 17.48 | 0 | 63.49 ± 17.48 | 0.003 |
| Age group, n(%) | 0 |  | 0 |  |  |
| 18-44 |  | 12,801 (15.8%) |  | 1,198,795 (14.9%) | 0.025 |
| 45-64 |  | 27,255 (33.7%) |  | 2,586,531 (32.1%) | 0.034 |
| 65-79 |  | 26,171 (32.4%) |  | 2,665,208 (33.1%) | 0.015 |
| 80-100 |  | 14,589 (18.1%) |  | 1,601,598 (19.9%) | 0.045 |
| Sex | 0 |  | 0 |  |  |
| Female |  | 43,448 (53.8%) |  | 4,302,754 (53.4%) | 0.008 |
| Male |  | 36,899 (45.7%) |  | 3,703,849 (46.0%) | 0.006 |
| Unspecified/Other |  | 469 (0.5%) |  | 45,539 (0.6%) | 0.013 |
| Ethnicity | 0 |  | 0 |  |  |
| Hispanic or Latino |  | 4,419 (5.5%) |  | 433,347 (5.4%) | 0.004 |
| Non-Hispanic or Latino |  | 73,433 (90.9%) |  | 7,323,876 (91.0%) | 0.003 |
| Unspecified/Other |  | 2,964 (3.7%) |  | 294,910 (3.7%) | 0.000 |
| Multiracial | 0 |  |  |  |  |
| Yes |  | 6,499 (8.0%) |  | 638,430 (7.9%) | 0.004 |
| Race (First Category) | 0 |  | 0 |  |  |
| White |  | 63,530 (78.6%) |  | 6,341,788 (78.8%) | 0.005 |
| Black or African American |  | 11,707 (14.5%) |  | 1,158,050 (14.4%) | 0.003 |
| Asian |  | 1,834 (2.3%) |  | 176,207 (2.2%) | 0.007 |
| American Indian or Alaska Native |  | 807 (1.0%) |  | 82,565 (1.0%) | 0.000 |
| Native Hawaiian or Other Pacific Islander |  | 287 (0.4%) |  | 29,054 (0.4%) | 0.000 |
| Unspecified/Other |  | 2,649 (3.2%) |  | 264,469 (3.3%) | 0.006 |
| Financial Class | 0 |  | 0 |  |  |
| Medicare |  | 32,899 (40.7%) |  | 3,278,868 (40.7%) | 0.000 |
| Medicaid |  | 5,906 (7.3%) |  | 589,793 (7.3%) | 0.000 |
| Miscellaneous/Other |  | 39,100 (48.3%) |  | 3,886,683 (48.3%) | 0.000 |
| Self-Pay |  | 1,228 (1.5%) |  | 128,393 (1.6%) | 0.008 |
| Unspecified |  | 1,628 (2.0%) |  | 168,396 (2.1%) | 0.007 |
| Primary RUCA | 644 |  | 65054 |  |  |
| Metropolitan |  | 66,256 (82.3%) |  | 6,632,316 (82.3%) | 0.000 |
| Micropolitan |  | 7,104 (8.8%) |  | 704,477 (8.7%) | 0.003 |
| Small town |  | 3,742 (4.6%) |  | 380,834 (4.7%) | 0.005 |
| Rural |  | 2,774 (3.4%) |  | 269,452 (3.3%) | 0.006 |
| Social Vulnerability Index Overall, mean (SD) | 828 | 0.57 ± 0.28 | 85889 | 0.57 ± 0.28 | 0.000 |
| Social Vulnerability Index Theme 1, mean (SD) | 828 | 0.51 ± 0.30 | 85889 | 0.51 ± 0.30 | 0.000 |
| Social Vulnerability Index Theme 2, mean (SD) | 828 | 0.57 ± 0.27 | 85889 | 0.57 ± 0.27 | 0.000 |
| Social Vulnerability Index Theme 3, mean (SD) | 828 | 0.61 ± 0.24 | 85889 | 0.61 ± 0.24 | 0.000 |
| Social Vulnerability Index Theme 4, mean (SD) | 828 | 0.60 ± 0.26 | 85889 | 0.60 ± 0.26 | 0.000 |
| **Outcome** |  |  |  |  |  |
| 1 Year (365 days) Mortality | 0 |  | 0 |  |  |
| Yes |  | 5,476 (6.8%) |  | 537,945 (6.7%) | 0.004 |
| 3 Year (1095 days) Mortality | 0 |  | 0 |  |  |
| Yes |  | 10,454 (13.0%) |  | 1,049,894 (13.0%) | 0.000 |
| 5 Year (1825 days) Mortality | 0 |  | 0 |  |  |
| Yes |  | 14,655 (18.2%) |  | 1,469,026 (18.2%) | 0.000 |
| Inpatient Length of Stay (days) |  | 4.19 ± 5.71 |  | 4.27 ± 5.72 | 0.014 |

SD, standard deviation; SMD, standardized mean difference; SVI, Social Vulnerability Index; RUCA, Rural-Urban Commuting Area. SneakPeek represents a pre-designated 1% random sample of Epic Cosmos records held out prior to model development and used as the test set for evaluation.

#### 2.2.2 Comorbidities

Comorbidities were identified using ICD-10 diagnosis codes from all diagnosis positions mapped to the AHRQ Clinical Classifications Software Refined (CCSR) for ICD-10-CM (2025 version).^20^ Two clinicians (RL, BTR) independently reviewed all 553 CCSR diagnostic categories and selected clinically relevant comorbid conditions. Only conditions meeting AHRQ chronic condition criteria (2025 version) were retained, defined as conditions lasting at least 12 months and either placing limitations on self-care or independent living, or requiring ongoing medical intervention.^21^

#### 2.2.3 Outcomes

The primary outcome was all-cause mortality at 1 year (365 days) following the index admission, ascertained using death dates recorded in Epic Cosmos. Epic Cosmos aggregates mortality information from multiple sources, capturing deaths beyond the hospital setting. Secondary outcomes were all-cause mortality at 3 and 5 years (1095 and 1825 days).

### 2.3 Personalized Comorbidity Score Development

PCS is intended for use as a summary covariate in downstream analyses, not as a standalone risk prediction tool. A Cox proportional hazards model serves as the underlying estimation framework to derive the score, and evaluation metrics are reported to characterize its performance properties relative to existing indices.

Epic Cosmos provides a pre-designated 1% random sample of records, referred to as SneakPeek, which was held out prior to model development. The remaining 99% of records were used for score development. The SneakPeek subset was not used for model fitting or tuning and was reserved solely for final evaluation in a held-out test set.

To account for differences in comorbidity patterns and risk across demographic groups, models were developed separately by age (18-44, 45-64, 65-79, and ≥80 years) and sex (male or female). Age groups were defined based on MeSH age categories.^22^ Two model specifications were considered: PCS Core, which included age and comorbidity conditions as predictors, and PCS Extended, which additionally incorporated socioeconomic and geographic context, including the SVI, RUCA classification, and insurance status.

Scores were estimated separately within each age subgroup. We prespecified evaluation of both age-stratified and age-unified specifications for each age group. The specification demonstrating superior discrimination in the held-out data was retained. This was done because younger age groups have relatively few outcome events, which could lead to unstable coefficient estimates.

#### 2.3.1 Two-Stage Cox Model Development

The comorbidity score was constructed using a two-stage framework consisting of variable selection followed by final model estimation. The dataset was randomly divided into two equal subsets: one used for selecting relevant predictors and the other used for fitting the final model. This was done to reduce selection bias because using the same data for both variable selection and coefficient estimation can yield inflated and unstable estimates.

##### 2.3.1.1 Feature Selection

Feature selection was performed using LASSO-penalized Cox proportional hazards regression applied to the first subset. Predictors with nonzero coefficients were retained to form the final set of selected covariates. The penalty parameter was selected via 5-fold cross-validation within the first subset.

##### 2.3.1.2 Final Model Estimation

The final model was estimated using a Cox proportional hazards model with the features selected in Stage 1, fitted on the second subset. Age was modeled using cubic B-spline basis functions to capture potential nonlinear relationships with risk. For an individual with covariate vector *X_i_*, the hazard function was modeled as

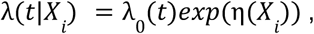

where λ_0_(*t*) is the baseline hazard and the risk score η(*X_i_*) is a linear combination of all binary predictors and B-spline basis transformations of age. Estimated coefficients with negative values for comorbidities were set to zero. Negative weights in comorbidity scoring systems have been noted as statistical artifacts that do not reflect genuinely protective effects on mortality.^23^ However, we acknowledge that some negative coefficients may reflect genuinely protective conditions or collinear features. A sensitivity analysis retaining negative coefficients was therefore conducted, and both the constrained and unconstrained model specifications are made available to allow researchers to select the version most appropriate for their use case.

#### 2.3.2 Translating the Linear Predictor into a Comorbidity Score

To provide a clinically interpretable measure of health risk, the linear predictor η(*X_i_*) derived from the fitted Cox model was translated into a comorbidity score using restricted mean survival time (RMST). For a prespecified time horizon τ, the RMST is defined as

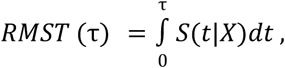

where *S*(*t*|*X*) is the predicted survival function for a patient with covariate vector *X*. Under the fitted Cox model, the predicted survival function is

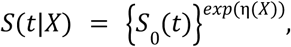

where *S*_0_(*t*) denotes the estimated baseline survival function. The comorbidity score was defined as the predicted RMST for each individual, rescaled and inverted so that higher values indicate greater comorbidity burden:

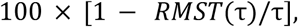

which can be interpreted as 100 times the average cumulative mortality rate over the interval[0, τ]. For example, a patient with a PCS score of 5 has an estimated average cumulative mortality rate of 5% over one year, while a patient with a score of 40 has an estimated rate of 40%.

While linear predictor η(*X_i_*) forms the basis of the score, its log-hazard scale is relative and requires a reference group to interpret directly. The RMST transformation converts this into an absolute, percentage-scale measure that is more intuitive for summarizing comorbidity burden across patients.

The Cox proportional hazards approach was selected in part because it effectively handles right censoring present in survival data, which is common in EHR-based cohorts. The two-stage approach was adopted with two objectives. The first stage screens informative predictors, as model sparsity was preferred to support reproducibility and generalizability. The second stage generates unbiased coefficient estimates for the effect of each comorbidity.

### 2.4 Descriptive statistics

Patient demographic characteristics and comorbidity prevalence were summarized for the full analytic cohort and the held-out SneakPeek 1% subset. Continuous variables were reported as means and standard deviations and categorical variables as counts and proportions. Standardized mean differences (SMD) were calculated to assess representativeness of the SneakPeek subset relative to the full cohort. The ten most prevalent CCSR-defined comorbidities were identified and reported. Score distributions were examined across indices. A LOWESS trend line was fitted to visualize the relationship between age at admission and each comorbidity score.

### 2.5 Model Evaluation

As a summary covariate designed for risk adjustment, PCS was evaluated on its ability to discriminate and characterize disease burden relative to existing indices, rather than as a standalone risk prediction tool. PCS was compared against three traditional indices: the original Charlson Comorbidity Index (Original CCI), the Quan-updated Charlson Comorbidity Index (Quan CCI), and the van Walraven Elixhauser Comorbidity Index (van Walraven ECI).^5,8,17^ These were selected as comparators given their widespread adoption in research practice and inclusion in commonly used comorbidity software package.^24,25^ Comorbidity conditions for each traditional index were identified using established ICD-10 coding algorithms.^7^ A consistent two-year lookback window was applied across all indices. No correction for multiple comparisons was applied, as automatic adjustment may increase type II error and is not warranted for empirical observations.^26^ Instead, 95% confidence intervals are reported to support interpretation of findings.

#### 2.5.1 Model performance

Score discrimination was evaluated as the primary performance metric. Area under the receiver operating characteristic curve (AUROC) was the primary discrimination metric, assessed in the held-out SneakPeek 1% dataset using AUROC for classifying death and survivor at given time points. The C-statistic, estimated using survival time to account for censoring, was reported as a complementary measure. Both metrics were evaluated with 95% confidence intervals derived from 2,000 bootstrap iterations in the held-out set. P-values below 0.001 cannot be precisely estimated given the number of bootstrap iterations.

Comorbidity scores are not inherently calibrated to predicted probabilities, and raw scores from different indices are on incomparable scales. To enable consistent comparison across all indices, each score was mapped to outcome probability using Platt scaling with 5-fold cross-validation in the training set.

Calibration curves are presented to characterize how each score tracks observed mortality rates across the risk spectrum. Specifically, each score was mapped to outcome probability using Platt scaling:

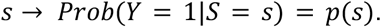

where *S* is the comorbidity score and *Y* is the binary event indicator, with the mapping estimated via 5-fold cross-validation in the training set. These derived mappings were then applied to the held-out dataset, where calibration was evaluated using decile-based curves comparing mean predicted probabilities against observed event rates. Calibration curves are presented to characterize how each score tracks observed mortality rates across the risk spectrum.

#### 2.5.2 Fairness analysis

Calibration and discrimination were compared across seven demographic subgroups as a measure of score fairness: age, sex, race, ethnicity, insurance class, SVI quartile, and RUCA classification. Race and ethnicity were not used in calculating PCS or the traditional indices but were evaluated as dimensions of fairness. While overall calibration is not a primary performance metric, subgroup-level calibration is prioritized as the primary fairness metric because systematic differences in calibration across demographic groups may differentially affect score performance for specific populations.^27^

Maximum calibration error (MCE) was the primary fairness metric, defined as

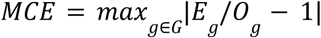

the maximum absolute difference between the expected-to-observed (E/O) ratio and 1.0 across demographic subgroups, capturing the worst-case disparity in calibration. Here, *E_g_* and *Og* represent the expected mortality rate in subgroup *g* according to the calibrated index and observed mortality rate.

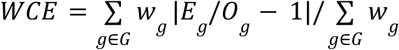

The weighted calibration error (WCE), the subgroup-size-weighted average of absolute calibration errors, was examined as a secondary fairness metric, where *w_g_* is the subgroup size.

Subgroup-level calibration curves are not presented for each demographic group, as some subgroups have insufficient sample sizes to support reliable decile-based calibration estimates. MCE and WCE are used instead to quantitatively summarize calibration differences across subgroups in a comparable format. Subgroup-level AUROC values were reported for each demographic group to confirm that PCS maintains higher discrimination than traditional indices within individual subgroups. AUROC range across subgroups was also computed as a complementary measure of discrimination consistency. Group fairness metrics based on classification thresholds were not applied, as they require a fixed decision threshold and preclude meaningful comparisons across indices with fundamentally different scoring systems.^27^ Subgroup-level confidence intervals were derived from 500 bootstrap iterations. P-values below 0.004 cannot be precisely estimated given the number of bootstrap iterations.

#### 2.5.3 Subgroup-Specific Condition Weights

To illustrate how comorbidity weights vary across demographic subgroups, log hazard ratios for each CCSR condition were visualized across all age-sex subgroups. This descriptive analysis highlights the degree to which individual conditions contribute differently to the score depending on a patient’s age and sex, supporting the rationale for subgroup-specific weighting.

### 2.6 Open-source Software Package Development

These packages enable researchers to compute PCS Core and PCS Extended scores directly from CCSR-coded diagnoses using pre-estimated condition weights. Users can specify the score variant (PCS Core or PCS Extended), outcome (1, 3, and 5 year mortality) and the package version. Each parameter selects the corresponding set of pre-estimated weights, allowing researchers to apply the appropriate model for their setting. Package versions will be updated periodically to reflect the evolving clinical landscape and refinements to the framework. Guidance on minimum data requirements and handling of missing or incomplete input data is provided in the package documentation.

### 2.7 Validation

We validated model performance in two complementary EHR datasets: Stanford EHR and MIMIC-IV hospital admission data^28^. Stanford EHR comprised comprehensive inpatient and outpatient data from Stanford Health Care, an academic medical center in Northern California. MIMIC-IV is a publicly available EHR dataset from Beth Israel Deaconess Medical Center, a tertiary academic medical center in Boston, Massachusetts. Neither dataset constitutes a fully external validation, as both source institutions participate in the Epic Cosmos network,^29^ introducing the possibility of patient overlap with the development cohort. Nevertheless, the two datasets differ meaningfully in patient populations, data structure, and institutional context, providing complementary evaluations of model performance.

The Epic Cosmos cohort design was largely replicated in Stanford EHR, given access to comprehensive inpatient and outpatient data. In contrast, MIMIC-IV contains inpatient data only and does not support full replication, including the fixed observation period, longitudinal outpatient encounters, and prior healthcare utilization requirements. For MIMIC-IV, the analytic cohort included non-pregnant adults (≥18 years) with a first inpatient admission. Comorbid conditions were ascertained from diagnosis codes recorded prior to or during the index admission, and one-year mortality was defined as death within 365 days of discharge.

Pre-trained PCS models were applied without retraining or refitting. Discrimination was assessed using AUROC for one-year mortality and compared with the Original CCI, Quan-adapted CCI, and Van Walraven ECI. Because MIMIC-IV lacked variables required for PCS Extended, only PCS Core was evaluated in that dataset.

### 2.8 Ethics Statement

This study was conducted in accordance with institutional guidelines. Stanford EHR data were used under an Institutional Review Board (IRB) approval, with a waiver of informed consent (Protocol 47644). Epic Cosmos data were determined to not constitute human subjects research by the Stanford Human Research Protection Program. MIMIC-IV is a publicly available, de-identified dataset exempt from IRB review.

## 3. Results

### 3.1 Study population

We identified a base inpatient cohort of 8,896,169 admissions from a static extract of Epic Cosmos (extract date: October 13, 2025), including patients with at least one hospital admission between March 31, 2015 and March 31, 2020. We sequentially excluded pregnancy-related admissions, same-day discharges, and encounters with insufficient continuity of care; patients aged under 18 or over 100 years; those not eligible for Cosmos Data Analytics; patients residing outside the United States; and records with invalid or incomplete data. The final analytic cohort comprised 8,052,133 admissions (Figure 2).

### 3.2 Cohort characteristics

The mean age was 63.5 years (SD 17.5), and females comprised 53.4% of the cohort. The majority of patients identified as non-Hispanic or Latino (91.0%) and White (78.8%). Medicare was the most common financial class (40.7%), and most patients resided in metropolitan areas (82.3%). The SVI Overall mean was 0.57 (SD 0.28). One-year, three-year, and five-year mortality rates were 6.7%, 13.0%, and 18.2%, respectively. The SneakPeek 1% dataset (n=80,816) used as the held-out test set showed similar demographic and outcome distributions across all characteristics, with SMD less than 0.05 (Table 1)

The ten most prevalent comorbidities in the full dataset were essential hypertension (54.7%), disorder of lipid metabolism (43.7%), and esophageal disorders (25.0%), followed by type 2 diabetes, osteoarthritis, spondylopathies, diabetes without complication, coronary atherosclerosis, obesity, and nervous system pain syndrome (18.7-25.0%; Figure S1).

### 3.3 PCS Evaluation

All indices showed right-skewed score distributions. PCS Core and PCS Extended scores increase more steeply across the age range (Figure S2 and S3).

#### 3.3.1 Model performance

PCS Core and PCS Extended outperformed all three traditional indices across all time horizons. For both the Core and Extended versions, the age-unified model was applied to the younger age group (18-44 years), consistent with the prespecified model selection strategy described in the Methods. Using our primary outcome of 1-year mortality, PCS Core and PCS Extended achieved AUROC of 0.812 (95% CI 0.805-0.817) and 0.813 (95% CI 0.807-0.818), respectively, compared to 0.730 (95% CI 0.723-0.737) for the van Walraven ECI, 0.719 (95% CI 0.712-0.726) for the Quan CCI, and 0.714 (95% CI 0.707–0.721) for the Original CCI (P<0.001; Figure 3). Similar patterns were observed at 3-year (PCS Core: 0.799, 95% CI 0.795-0.803; PCS Extended: 0.800, 95% CI 0.795–0.804) and 5-year horizons (PCS Core: 0.795, 95% CI 0.790-0.799; PCS Extended: 0.796, 95% CI 0.792-0.800), consistently exceeding traditional indices across all time points (Table S2-S4). A sensitivity analysis retaining negative coefficients yielded slightly higher discrimination, suggesting that the constraint had minimal impact on performance (Table S2-S4).

**Figure 3.**
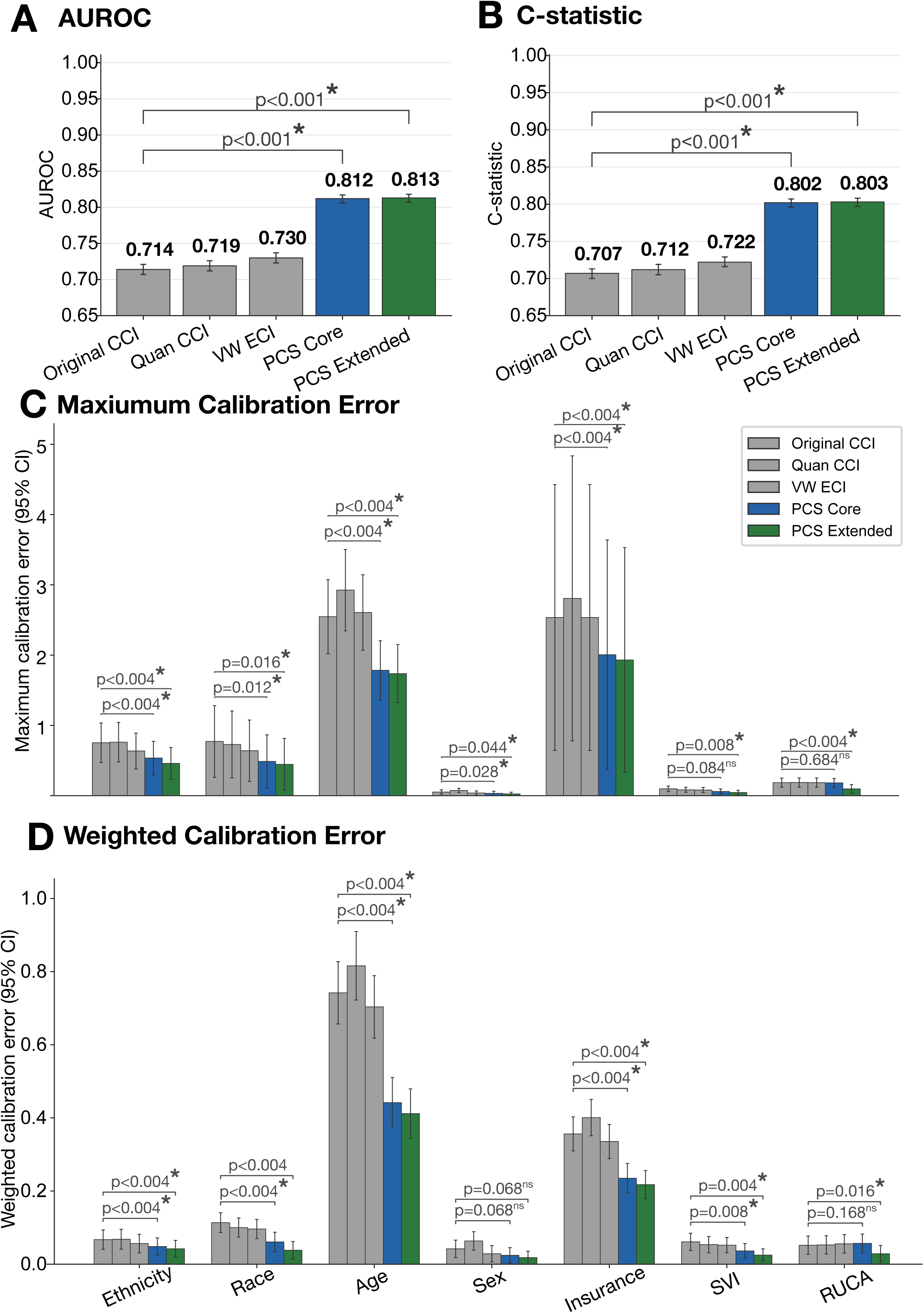
Performance comparison of the Personalized Comorbidity Score (PCS) and traditional comorbidity indices for one-year mortality prediction in the Epic Cosmos development cohort. Abbreviations: AUROC, area under the receiver operating characteristic curve; CCI, Charlson Comorbidity Index; PCS, Personalized Comorbidity Score; VW ECI, van Walraven Elixhauser Index; SVI, Social Vulnerability Index; RUCA, Rural-Urban Commuting Area. (A) AUROC. (B) Harrell’s C-statistic. (C) Maximum calibration error across demographic and socioeconomic subgroups. (D) Weighted calibration error across demographic and socioeconomic subgroups. Performance of the Original Charlson Comorbidity Index (CCI), Quan CCI, van Walraven Elixhauser Index (VW ECI), PCS Core, and PCS Extended are shown. Error bars represent 95% confidence intervals. Statistical significance in panels A and B was assessed using bootstrap resampling with 2,000 resamples; therefore, the smallest observable two-sided *P* value was 0.001. Statistical significance in panels C and D was assessed using bootstrap resampling with 500 resamples; therefore, the smallest observable two-sided *P* value was 0.004. Asterisks indicate statistically significant differences (*P* < 0.05), and “ns” indicates not statistically significant.

PCS Core and PCS Extended demonstrated wider predicted probability ranges than traditional indices, reflecting stronger risk differentiation consistent with higher discrimination. Both PCS variants showed systematic underestimation of risk at lower predicted probabilities, with observed event rates consistently exceeding predicted values in that range (Figure S4, S5, and S6).

#### 3.3.2 Fairness analysis

Calibration and discrimination were compared across all demographic subgroups, with both PCS variants showing reduced calibration error compared to traditional indices. MCE and WCE were consistently lower for PCS Core and PCS Extended across all subgroups (Table 2 and Figure 3). The direction and relative magnitude of calibration differences across subgroups are illustrated in Figure S7, which is presented to show directionality rather than inferential comparisons.

**Table 2.** Fairness metrics across demographic and socioeconomic subgroups.

| Model | Max Calibration Error (95% CI); P value | Weighted Calibration Error (95% CI); P value | AUROC Range (95% CI); P value | C-stat Range (95% CI); P value |
| --- | --- | --- | --- | --- |
| **Ethnicity** |  |  |  |  |
| Original CCI (ref) | 0.752 (0.494-1.058) | 0.067 (0.042-0.095) | 0.065 (0.026-0.101) | 0.067 (0.030-0.102) |
| Quan CCI | 0.761 (0.506-1.068) | 0.068 (0.042-0.096) | 0.059 (0.019-0.098) | 0.061 (0.021-0.099) |
| VW ECI | 0.636 (0.403-0.912) | 0.056 (0.034-0.084) | 0.057 (0.021-0.092) | 0.059 (0.023-0.093) |
| PCS Core | 0.535 (0.316-0.791); p<0.004 | 0.048 (0.026-0.073); p<0.004 | 0.035 (0.005-0.068); p=0.112 | 0.038 (0.007-0.070); p=0.108 |
| PCS Extended | 0.459 (0.249-0.700); p<0.004 | 0.042 (0.021-0.067); p<0.004 | 0.034 (0.003-0.068); p=0.100 | 0.038 (0.006-0.070); p=0.104 |
| **Race** |  |  |  |  |
| Original CCI (ref) | 0.772 (0.390-1.410) | 0.114 (0.087-0.141) | 0.107 (0.055-0.177) | 0.108 (0.056-0.176) |
| Quan CCI | 0.729 (0.368-1.320) | 0.100 (0.075-0.127) | 0.114 (0.054-0.200) | 0.114 (0.055-0.197) |
| VW ECI | 0.640 (0.311-1.185) | 0.096 (0.072-0.124) | 0.122 (0.053-0.254) | 0.123 (0.054-0.250) |
| PCS Core | 0.488 (0.208-0.963);<br>p=0.012 | 0.061 (0.036-0.088);<br>p<0.004 | 0.109 (0.056-0.211);<br>p=0.848 | 0.106 (0.056-0.200);<br>p=0.860 |
| PCS Extended | 0.446 (0.177-0.914);<br>p=0.016 | 0.038 (0.017-0.064);<br>p<0.004 | 0.112 (0.055-0.221);<br>p=0.804 | 0.109 (0.056-0.208);<br>p=0.848 |
| **Age** |  |  |  |  |
| Original CCI<br>(ref) | 2.547 (2.070-3.122) | 0.742 (0.663-0.834) | 0.182 (0.143-0.226) | 0.190 (0.152-0.232) |
| Quan CCI | 2.925 (2.402-3.557) | 0.816 (0.730-0.917) | 0.166 (0.127-0.208) | 0.175 (0.135-0.215) |
| VW ECI | 2.606 (2.111-3.184) | 0.704 (0.625-0.796) | 0.167 (0.139-0.206) | 0.174 (0.147-0.214) |
| PCS Core | 1.784 (1.395-2.239);<br>p<0.004 | 0.442 (0.377-0.515);<br>p<0.004 | 0.128 (0.100-0.163);<br>p<0.004 | 0.141 (0.112-0.176);<br>p=0.008 |
| PCS Extended | 1.737 (1.357-2.184);<br>p<0.004 | 0.412 (0.349-0.484);<br>p<0.004 | 0.117 (0.092-0.1492);<br>p<0.004 | 0.130 (0.103-0.162);<br>p<0.004 |
| **Sex** |  |  |  |  |
| Original CCI<br>(ref) | 0.052 (0.023-0.084) | 0.042 (0.019-0.067) | 0.011 (0.001-0.025) | 0.010 (0.001-0.023) |
| Quan CCI | 0.074 (0.046-0.106) | 0.064 (0.038-0.089) | 0.013 (0.001-0.028) | 0.012 (0.001-0.026) |
| VW ECI | 0.038 (0.010-0.069) | 0.029 (0.007-0.052) | 0.012 (0.001-0.025) | 0.010 (0.001-0.022) |
| PCS Core | 0.033 (0.006-0.064);<br>p=0.028 | 0.024 (0.005-0.047);<br>p=0.068 | 0.010 (0.001-0.021);<br>p=0.948 | 0.011 (0.001-0.023);<br>p=0.896 |
| PCS Extended | 0.025 (0.004-0.053);<br>p=0.044 | 0.018 (0.003-0.038);<br>p=0.068 | 0.011 (0.001-0.022);<br>p=1.000 | 0.0112 (0.001-0.023);<br>p=0.796 |
| **Insurance** |  |  |  |  |
| Original CCI<br>(ref) | 2.536 (0.517-4.300) | 0.356 (0.311-0.404) | 0.134 (0.087-0.240) | 0.138 (0.090-0.246) |
| Quan CCI | 2.807 (0.614-4.670) | 0.401 (0.352-0.452) | 0.128 (0.071-0.241) | 0.134 (0.074-0.248) |
| VW ECI | 2.535 (0.509-4.291) | 0.336 (0.289-0.383) | 0.143 (0.084-0.242) | 0.149 (0.088-0.249) |
| PCS Core | 2.006 (0.247-3.515);<br>p<0.004 | 0.235 (0.196-0.277);<br>p<0.004 | 0.123 (0.089-0.208);<br>p=0.848 | 0.128 (0.094-0.212);<br>p=0.860 |
| PCS Extended | 1.931 (0.225-3.422);<br>p<0.004 | 0.218 (0.180-0.257);<br>p<0.004 | 0.123 (0.086-0.209);<br>p=0.804 | 0.127 (0.092-0.212);<br>p=0.848 |
| **SVI** |  |  |  |  |
| Original CCI<br>(ref) | 0.097 (0.060-0.140) | 0.061 (0.040-0.087) | 0.036 (0.018-0.054) | 0.035<br>(0.017-0.052) |
| Quan CCI | 0.083 (0.050-0.122) | 0.053 (0.033-0.077) | 0.036 (0.017-0.053) | 0.034 (0.016-0.051) |
| VW ECI | 0.081 (0.047-0.119) | 0.052 (0.031-0.073) | 0.025 (0.009-0.043) | 0.024 (0.009-0.042) |
| PCS Core | 0.060 (0.029-0.099);<br>p=0.084 | 0.036 (0.016-0.057);<br>p=0.008 | 0.028 (0.015-0.040);<br>p=0.316 | 0.027 (0.014-0.038)<br>p=0.288 |
| PCS Extended | 0.045 (0.016-0.081);<br>p=0.008 | 0.025 (0.010-0.044);<br>p=0.004 | 0.029 (0.015-0.042);<br>p=0.396 | 0.027 (0.015-0.040)<br>p=0.348 |
| **RUCA** |  |  |  |  |
| Original CCI<br>(ref) | 0.185 (0.118-0.247) | 0.052 (0.029-0.079) | 0.022 (0.004-0.048) | 0.021 (0.003-0.047) |
| Quan CCI | 0.188 (0.119-0.249) | 0.053 (0.029-0.079) | 0.022 (0.004-0.048) | 0.022 (0.003-0.047) |
| VW ECI | 0.186 (0.117-0.248) | 0.056 (0.032-0.082) | 0.034 (0.011-0.068) | 0.037 (0.011-0.066) |
| PCS Core | 0.182 (0.118-0.243);<br>p=0.684 | 0.057 (0.033-0.084);<br>p=0.168 | 0.016 (0.003-0.035);<br>p=0.848 | 0.016 (0.003-0.035);<br>p=0.860 |
| PCS Extended | 0.097 (0.039-0.160);<br>p<0.004 | 0.029 (0.009-0.054);<br>p=0.016 | 0.016 (0.003-0.037);<br>p=0.804 | 0.016 (0.003-0.037);<br>p=0.848 |

CI, confidence interval; CCI, Charlson Comorbidity Index; VW ECI, van Walraven Elixhauser Comorbidity Index; MCE, maximum calibration error; WCE, weighted calibration error; AUROC, area under the receiver operating characteristic curve; C-stat, concordance statistic. Original CCI serves as the reference group for comparison. Confidence intervals and p-values were derived from 500 bootstrap iterations; p-values below 0.004 cannot be precisely estimated given the number of iterations.

Age exhibited the largest calibration differences across all models. Traditional indices showed MCE ranging from 2.547 to 2.925, whereas PCS Core and PCS Extended reduced MCE to 1.784 (95% CI 1.395-2.239; P<0.004) and 1.737 (95% CI 1.358-2.184; P<0.004), respectively. In contrast, sex demonstrated the smallest calibration differences across all models.

Calibration improvements were also observed across race and ethnicity. For race, MCE was reduced to 0.488 (95% CI 0.208-0.963; *P*=0.012) for PCS Core and 0.446 (95% CI 0.177-0.914; *P*=0.012) for PCS Extended, compared with 0.640-0.772 for traditional indices. For ethnicity, MCE was reduced to 0.535 (95% CI 0.316-0.791; *P*<0.004) for PCS Core and 0.459 (95% CI 0.249-0.700; *P*<0.004) for PCS Extended, compared with 0.636-0.761 for traditional indices.

Insurance class also showed lower calibration error for both PCS variants. MCE was reduced to 2.006 (95% CI 0.247-3.515; *P*<0.004) for PCS Core and 1.931 (95% CI 0.225-3.422; *P*<0.004) for PCS Extended, compared with 2.535-2.807 for traditional indices.

For SVI and RUCA, PCS Extended demonstrated the largest reductions in calibration error. For SVI, MCE was reduced to 0.045 (95% CI 0.016-0.081; *P*=0.008) for PCS Extended, compared with 0.081-0.097 for traditional indices, whereas PCS Core showed an MCE of 0.060 (95% CI 0.029-0.099; *P*=0.084). For RUCA, MCE was reduced to 0.097 (95% CI 0.039-0.160; *P*<0.004) for PCS Extended, compared with 0.185-0.188 for traditional indices, whereas PCS Core showed minimal improvement (0.182, 95% CI 0.118-0.243; *P*=0.684).

Subgroup-level AUROC was consistently higher for PCS Core and PCS Extended than for traditional indices across all subgroup definitions, while the range of AUROC across subgroups remained comparable between PCS variants and traditional indices (Table 2, Figure S8, and Figure S9).

#### 3.3.3 Subgroup-Specific Condition Weights

From an initial pool of 248 clinician-selected CCSR comorbidities, PCS Core selected 120 unique CCSR comorbidity features across all age-sex subgroups, with the number of features per subgroup ranging from 58 to 97. PCS Extended selected 152 unique features, ranging from 89 to 130 per subgroup. Figure S10 illustrates subgroup-specific weighting across all CCSR disease categories, with neoplasm and malignancy categories showing the most pronounced variation. Secondary malignancies (NEO070, metastatic or secondary malignancy) had the highest mean absolute coefficient in both PCS variants. In PCS Core, its coefficient ranged from 1.549 in females aged 45-64 to 0.743 in males aged 80 and older. In PCS Extended, it ranged from 1.563 in females aged 45-64 to 0.743 in males aged 80 and older.

### 3.5 Open-source software package

Open-source R and Python packages have been developed and will be made publicly available upon journal acceptance (https://github.com/su-boussard-lab/personalized-comorbidity-score). Package versions will be updated periodically to reflect the evolving clinical landscape and refinements to the framework.

### 3.6 Validation

We evaluated PCS in two complementary EHR datasets with distinct patient populations and data structures: Stanford Health Care and MIMIC-IV. Descriptive characteristics of both validation cohorts are summarized in Tables S5 and S6. In the Stanford Health Care cohort, PCS Core and PCS Extended achieved AUROCs of 0.778 and 0.777 for one-year mortality, respectively, outperforming the Original CCI (0.648), Quan CCI (0.654), and van Walraven ECI (0.653).

In the MIMIC-IV cohort, PCS Core achieved the highest discrimination for one-year mortality, with an AUROC of 0.847. In comparison, the van Walraven ECI achieved an AUROC of 0.807, followed by the Quan CCI (0.787) and the Original CCI (0.785).

## 4. Discussion

Traditional comorbidity indices treat disease burden as fixed and universal, yet the prognostic significance of comorbid conditions varies by age, sex, socioeconomic context, and the outcome being measured. Comorbidity scores are often used as generic control variables in research models. Because these indices apply uniform weights regardless of population, the resulting score may not accurately reflect the disease burden of the specific population being studied. PCS was designed to treat comorbidity burden as context-dependent and population-specific. It was built from over 8 million adult inpatient encounters across the Epic Cosmos network. PCS was designed to address four longstanding limitations of traditional indices: fixed comorbidity weights that fail to reflect heterogeneous disease burden, incomplete condition coverage, lack of socioeconomic context where available, and the absence of a systematic updating process. Both PCS variants showed numerically lower calibration error than traditional indices across all demographic subgroups examined, though this difference reached statistical significance in most but not all subgroups. These gains were achieved without including race or ethnicity as model features, which avoids the risk of reinforcing race-based disparities in research scoring.

PCS achieved significantly higher discrimination than traditional indices for 1-year all-cause mortality prediction (P<0.001). Prior efforts to improve comorbidity measurement through updates, adaptations, and machine learning approaches have achieved comparable or higher discrimination in some settings. ^1,2,6–8,13,30–34^ However, to our knowledge, no widely adopted comorbidity framework has simultaneously addressed comprehensive condition coverage, socioeconomic context, and demographic-stratified weighting.

PCS showed reduced subgroup calibration error across all demographic subgroups without including race or ethnicity as model features. Encoding race and ethnicity directly in comorbidity measurement has raised concerns about reinforcing race-based disparities in how disease burden is quantified and applied.^35,36^ PCS achieved improved subgroup calibration through age-sex stratified comorbidity selection and weighting. This contrasts with Tangel et al., who derived race/ethnicity-specific weights from binary Elixhauser conditions.^37^ Subgroup-level AUROC was consistently higher for both PCS variants compared to traditional indices across all demographic groups, indicating that the discrimination advantage of PCS is maintained within individual subgroups. Unlike traditional indices that apply uniform weights regardless of population, PCS provides comorbidity weights tailored to the age and sex profile of the study population, offering researchers a more meaningful measure of disease burden for their specific cohort. Age showed the largest absolute MCE improvement, reflecting the heterogeneity in disease burden across the lifespan that uniform-weight indices fail to capture.

PCS showed greater miscalibration than traditional indices at lower predicted probabilities, with observed event rates exceeding predicted values in that range. A known trade-off exists between discrimination and calibration in prediction modeling, where stronger discrimination can come at the cost of calibration.^38,39^ While calibration is critical for standalone clinical prediction tools where absolute risk estimates inform patient decisions,^27,38^ PCS is designed for use as a summary covariate for risk adjustment and as an input feature in downstream models. In both contexts, the primary criterion is discrimination, as the utility of PCS lies in correctly ranking patients by disease burden. Researchers should nonetheless consider this miscalibration pattern when interpreting absolute probability estimates derived from PCS at the lower end of the risk spectrum.

Discrimination was nearly identical between PCS Core and PCS Extended despite the latter incorporating additional socioeconomic and geographic features. This suggests that age-sex stratified comorbidity weighting alone captures much of the predictive signal for 1-year mortality. Alternatively, socioeconomic features may contribute more meaningfully to outcomes closely tied to social determinants of health, such as readmission.^40^ PCS Extended demonstrated superior calibration fairness for insurance class, SVI, and RUCA, consistent with its incorporation of socioeconomic features. For settings where socioeconomic data are available, PCS Extended may be preferable given its improved fairness profile. Where such data are unavailable, PCS Core remains a strong alternative with improved performance and fairness over traditional indices.

PCS assigns comorbidity weights that vary by age-sex subgroup, reflecting the clinical reality that the same condition carries different prognostic weight depending on a patient’s age and sex.^41–43^ Secondary malignancies showed the highest mean absolute coefficient among all features in both PCS variants, yet its weight was markedly lower in patients aged 80 and older compared to middle-aged patients. This reflects the diminishing relative contribution of malignancy to mortality risk in older patients, who carry a higher burden of competing causes of death.^44^ These patterns illustrate why a single set of comorbidity weights applied uniformly across all patients may systematically misrepresent disease burden in specific subgroups.

Discrimination in the 18-44 age group was lower when using fully age-stratified coefficients, likely reflecting the relatively low mortality rate in younger patients and the limited number of outcome events available for stable estimation. To address this, we evaluated both age-stratified and age-unified model specifications for each age group, adopting the specification with better predictive performance. This highlights an important practical consideration in personalized comorbidity frameworks. Subgroup-specific modeling requires sufficient outcome events, and excessive stratification in low-event subgroups may reduce rather than improve model performance.

PCS maintained performance across two distinct EHR datasets. However, neither constitutes a fully external validation given both institutions’ participation in the Epic Cosmos network. Although PCS consistently outperformed traditional comorbidity indices in both datasets, the magnitude of improvement was smaller in MIMIC-IV than in Stanford Health Care and the Epic Cosmos test cohort. Several factors may explain this finding. First, MIMIC-IV is an inpatient-only database that does not support reconstruction of the longitudinal outpatient history used during PCS development. This limits the availability of chronic disease information. Second, to ensure adequate comorbidity ascertainment in MIMIC-IV, diagnoses recorded during the hospitalization were included in score calculation, and outcome was measured from discharge rather than admission. In contrast, the Epic Cosmos and Stanford cohorts measured mortality from admission and used diagnoses recorded prior to admission only. These differences in diagnosis timing and outcome ascertainment likely improve discrimination for all comorbidity indices in MIMIC-IV, reducing the relative advantage of PCS. Despite these differences, PCS remained the best-performing comorbidity index across both validation datasets, supporting the robustness and generalizability of the framework.

This study has several notable strengths. PCS was developed on Epic Cosmos, the largest EHR database in the United States, including more than 8 million qualifying inpatient encounters across a geographically diverse network of participating health systems and payer types. Unlike traditional indices that rely on expert-curated condition lists with fixed weights, PCS uses data-driven condition selection and subgroup-specific weight estimation, allowing both the relevant conditions and their prognostic contributions to vary across age-sex subgroups. The use of AHRQ-defined CCSR categories and chronic condition indicators ensures comprehensive and standardized condition coverage through actively maintained and publicly available mappings, reducing the risk of variable selection bias inherent in expert-curated condition lists, while enabling a reproducible and updatable workflow. The framework is designed to support systematic weight updates as the clinical landscape evolves, addressing a key limitation of existing indices where updates become fragmented over time. The RMST-based scoring expresses comorbidity burden as an estimated cumulative mortality rate over the specified time horizon, offering a more interpretable scale than the log-hazard based linear predictor. The availability of versioned open-source R and Python packages supports reproducibility and broad adoption. Finally, the availability of both a core and extended version provides flexibility across research contexts, with PCS Core applicable where only clinical data are available and PCS Extended offering improved fairness where socioeconomic data can be linked.

This study has several limitations. First, although Epic Cosmos is broadly representative of the US population, it predominantly captures patients who receive care within the Epic network, potentially underrepresenting uninsured patients and those treated at non-Epic facilities such as the Veterans Health Administration. Second, PCS was developed and evaluated within the US healthcare context, and generalizability to other countries with distinct coding practices and care-seeking behaviors remains to be established. Third, while Epic Cosmos benefits from multi-source mortality ascertainment, EHR-based mortality capture remains inherently incomplete compared to national death registries, and death ascertainment practices vary across care sites, which may introduce differential outcome misclassification. Fourth, although PCS was validated in two independent datasets, Stanford Health Care and MIMIC-IV, neither constitutes a fully external validation, as both institutions participate in the Epic Cosmos network. True external validation in healthcare systems outside the Epic ecosystem remains an important next step before broad adoption. Fifth, incorporating socioeconomic variables such as SVI and insurance status may improve calibration across demographic subgroups but does not resolve underlying structural inequities and may encode existing disparities into the score.Finally, PCS was optimized for 1-year all-cause mortality, and extension to other outcomes would require outcome-specific retraining of the framework. In addition, the pre-trained weights provided reflect the Epic Cosmos population, and direct application to substantially different populations without validation may not be appropriate. Unlike traditional indices where fixed weights are applied universally, PCS is designed to be retrained on population-specific data, offering a path to address this limitation in future applications.

PCS represents a step toward comorbidity measurement that is data-driven, context-dependent, and extensible. Future work should establish best practices for adapting the PCS framework to new populations and outcomes, including guidance on minimum sample size requirements, cohort construction, retraining procedures, and version management. Beyond mortality, we hope to extend PCS to broader dimensions of health burden, including functional decline, quality of life, and healthcare utilization, each requiring outcome-specific retraining of the framework. PCS is designed to evolve alongside both clinical practice and advances in computational methods, with versioned weight updates and support for more complex modeling approaches as they become validated for this use case. Open-source R and Python packages are publicly available to support broad adoption. We envision an extensible design that could allow researchers and clinicians to contribute cohort-specific and outcome-specific variants as the framework grows, ultimately providing a foundation for precision public health research.^45^

## 5. Conclusion

Comorbidity measurement has long relied on indices developed decades ago, on limited populations, with fixed weights that do not reflect the diversity of patients seen in modern clinical practice. PCS illustrates a framework for modern comorbidity measurement that is data-driven, context-dependent, and designed to evolve. Across a nationally diverse cohort of over 8 million inpatient encounters, PCS showed higher discrimination than traditional indices and reduced calibration error across most demographic subgroups, without encoding race or ethnicity into the scoring model. As clinical research increasingly demands tools that are both accurate and population-specific, PCS provides a foundation for comorbidity measurement that is fit for the diversity of populations it serves. Unlike traditional indices with fixed weights applied universally, PCS is designed to be retrained and refined as populations, outcomes, and clinical contexts evolve, offering a systematic and extensible alternative to the fragmented updating that has long characterized comorbidity measurement.

## Data Availability

Epic Cosmos and Stanford Health Care EHR data contain protected health information and cannot be made publicly available due to patient privacy restrictions. Researchers may request access to Epic Cosmos through Epic’s Cosmos research program, subject to data use agreements. Stanford EHR data access requires institutional approval. MIMIC-IV is publicly available to credentialed users through PhysioNet (https://physionet.org/content/mimiciv/).

## Author Contributions

Y-M.H.: conceptualization, methodology, software, validation, formal analysis, investigation, resources, data curation, writing - original draft, writing - review & editing, visualization, project administration, funding acquisition; Y.C.: methodology, software, validation, formal analysis, investigation, resources, writing - original draft, writing - review & editing; J.X.: conceptualization, funding acquisition, writing - review & editing; T.P.: methodology, writing - review and editing; R.L.: validation, writing - review & editing; B.T.R.: validation, writing - review & editing. L.T.: conceptualization, methodology, formal analysis, investigation, supervision, writing: review and editing, funding acquisition. T.H-B.: conceptualization, resources, supervision, funding acquisition, writing - review and editing.

## Reference

1. Gagne, J. J., Glynn, R. J., Avorn, J., Levin, R. & Schneeweiss, S. A combined comorbidity score predicted mortality in elderly patients better than existing scores. J. Clin. Epidemiol. 64, 749–759 (2011).

2. De, G. V., Beckerman, H., Lankhorst, G. J. & Bouter, L. M. How to measure comorbidity: a critical review of available methods. Journal of clinical epidemiology 56, 221–229 (2003).

3. Austin, S. R., Wong, Y.-N., Uzzo, R. G., Beck, J. R. & Egleston, B. L. Why summary comorbidity measures such as the Charlson Comorbidity Index and Elixhauser score work. Med. Care 53, e65–72 (2015).

4. Soos, B. et al. Considerations for selecting and implementing comorbidity indices when using secondary data sources: a guide for health researchers. Int. J. Popul. Data Sci. 10, (2025).

5. Charlson, M. E., Pompei, P., Ales, K. L. & MacKenzie, C. R. A new method of classifying prognostic comorbidity in longitudinal studies: development and validation. J. Chronic Dis. 40, 373–383 (1987).

6. Charlson, M. E., Carrozzino, D., Guidi, J. & Patierno, C. Charlson Comorbidity Index: A critical review of clinimetric properties. Psychother. Psychosom. 91, 8–35 (2022).

7. Quan, H. et al. Coding algorithms for defining comorbidities in ICD-9-CM and ICD-10 administrative data. Med. Care 43, 1130–1139 (2005).

8. van Walraven, C., Austin, P. C., Jennings, A., Quan, H. & Forster, A. J. A modification of the Elixhauser comorbidity measures into a point system for hospital death using administrative data. Med. Care 47, 626–633 (2009).

9. Elixhauser, A., Steiner, C., Harris, D. R. & Coffey, R. M. Comorbidity measures for use with administrative data. Med. Care 36, 8–27 (1998).

10. NCI Comorbidity Index Overview. https://healthcaredelivery.cancer.gov/seermedicare/considerations/comorbidity.html.

11. Bateman, B. T. et al. Development of a comorbidity index for use in obstetric patients. Obstet. Gynecol. 122, 957–965 (2013).

12. England, B. R., Sayles, H., Mikuls, T. R., Johnson, D. S. & Michaud, K. Validation of the rheumatic disease comorbidity index: RDCI validation. Arthritis Care Res. (Hoboken) 67, 865–872 (2015).

13. Moore, B. J., White, S., Washington, R., Coenen, N. & Elixhauser, A. Identifying increased risk of readmission and in-hospital mortality using hospital administrative data: The AHRQ Elixhauser Comorbidity index: The AHRQ elixhauser comorbidity index. Med. Care 55, 698–705 (2017).

14. Nowels, M. A. & VanderWielen, L. M. Comorbidity indices: a call for the integration of physical and mental health. Prim. Health Care Res. Dev. 19, 96–98 (2018).

15. Pfaff, E. R. et al. Coding long COVID: characterizing a new disease through an ICD-10 lens. BMC Med. 21, 58 (2023).

16. Social determinants of health. https://www.who.int/health-topics/social-determinants-of-health#tab=tab_1.

17. Quan, H. et al. Updating and validating the Charlson comorbidity index and score for risk adjustment in hospital discharge abstracts using data from 6 countries. Am. J. Epidemiol. 173, 676–682 (2011).

18. Epic Cosmos. https://cosmos.epic.com/about/.

19. Noel, A. & Bartelt, K. Cosmos: Real-world data powered by the healthcare community. J. Soc. Clin. Data Manag. 3, (2023).

20. Clinical Classifications Software Refined (CCSR) for ICD-10-CM Diagnoses. https://hcup-us.ahrq.gov/toolssoftware/ccsr/dxccsr.jsp.

21. Chronic Condition Indicator Refined (CCIR) for ICD-10-CM. https://hcup-us.ahrq.gov/toolssoftware/chronic_icd10/chronic_icd10.jsp.

22. Adult - MeSH - NCBI. https://www-ncbi-nlm-nih-gov.stanford.idm.oclc.org/mesh/68000328.

23. Stausberg, J. & Hagn, S. New morbidity and comorbidity scores based on the structure of the ICD-10. PLoS One 10, e0143365 (2015).

24. comorbidipy. PyPI https://pypi.org/project/comorbidipy/.

25. Gasparini, A. Comorbidity: An R Package for Computing Comorbidity Scores. (Github).

26. Rothman, K. J. No adjustments are needed for multiple comparisons. Epidemiology 1, 43–46 (1990).

27. van der Meijden, S. L. et al. Navigating fairness in AI-based prediction models: Theoretical constructs and practical applications. medRxiv 2025.03.24.25324500 (2025) doi:10.1101/2025.03.24.25324500.

28. Johnson, A. E. W. et al. MIMIC-IV, a freely accessible electronic health record dataset. Sci. Data 10, 1 (2023).

29. Epic Cosmos. https://cosmos.epic.com/community/.

30. Park, J.-H. & Lim, J. A machine learning model for predicting severity-adjusted in-hospital mortality in pneumonia patients. *Digit*. Health 11, 20552076251351467 (2025).

31. Uddin, S. et al. Comorbidity and multimorbidity prediction of major chronic diseases using machine learning and network analytics. Expert Syst. Appl. 205, 117761 (2022).

32. Liu, J. et al. Using machine-learning methods to predict in-hospital mortality through the Elixhauser index: A Medicare data analysis. Res. Nurs. Health 46, 411–424 (2023).

33. Hyer, J. M. et al. Can we improve prediction of adverse surgical outcomes? Development of a surgical Complexity Score using a novel machine learning technique. J. Am. Coll. Surg. 230, 43–52.e1 (2020).

34. Chervu, N. L. et al. Development of a surgery-specific comorbidity score for use in administrative data. Ann. Surg. 10.1097/SLA.0000000000006544 (2024).

35. Churchwell, K., et al. Call to action: Structural racism as a fundamental driver of health disparities: A presidential advisory from the American Heart Association. Circulation 142, e454–e468 (2020).

36. Vyas, D. A., Eisenstein, L. G. & Jones, D. S. Hidden in Plain Sight — Reconsidering the Use of Race Correction in Clinical Algorithms. New England Journal of Medicine (2020) doi:10.1056/nejmms2004740.

37. Tangel, V. E., Lui, B., Halawani Aladdin, D. E., Pryor, K. O. & White, R. S. Validity of comorbidity adjustment scores in estimating in-hospital mortality in individual subgroups of race/ethnicity. J. Comp. Eff. Res. 10, 823–829 (2021).

38. Van Calster, B. et al. Calibration: the Achilles heel of predictive analytics. BMC Med. 17, 230 (2019).

39. Cook, N. R. Use and misuse of the receiver operating characteristic curve in risk prediction. Circulation 115, 928–935 (2007).

40. Zhang, Y. et al. Assessing the impact of social determinants of health on predictive models for potentially avoidable 30-day readmission or death. PLoS One 15, e0235064 (2020).

41. Mesas, A. E. et al. Predictors of in-hospital COVID-19 mortality: A comprehensive systematic review and meta-analysis exploring differences by age, sex and health conditions. PLoS One 15, e0241742 (2020).

42. Kartoun, U. et al. Prediction performance and fairness heterogeneity in cardiovascular risk models. Sci. Rep. 12, 12542 (2022).

43. Li, G. et al. The age-specific comorbidity burden of mild cognitive impairment: a US claims database study. Alzheimers. Res. Ther. 15, 211 (2023).

44. Daskivich, T. J. et al. Effect of age, tumor risk, and comorbidity on competing risks for survival in a U.S. population-based cohort of men with prostate cancer. Ann. Intern. Med. 158, 709–717 (2013).

45. Roberts, M. C., Holt, K. E., Del Fiol, G., Baccarelli, A. A. & Allen, C. G. Precision public health in the era of genomics and big data. Nat. Med. 30, 1865–1873 (2024).

