## Supplementary Material for "One Size Does Not Fit All: A Data-Driven Framework for Personalized Comorbidity Scoring"

**Supplementary Materials**

### **Supplementary Tables**

**Table S1.** Variable definitions

**Table S2.** Model discrimination for 1-year mortality

**Table S3.** Model discrimination for 3-year mortality

**Table S4.** Model discrimination for 5-year mortality

**Table S5.** Characteristics of the Stanford Health Care validation cohort

**Table S6.** Characteristics of the MIMIC-IV validation cohort

### **Supplementary Figures**

**Figure S1.** Ten most prevalent CCSR-defined chronic comorbidities in the Epic Cosmos cohort

**Figure S2.** Score distributions for traditional comorbidity indices and one-year PCS variants in the SneakPeek 1% held-out validation dataset

**Figure S3.** Relationship between age and comorbidity scores (LOWESS curves)

**Figure S4.** Calibration curves for one-year mortality across comorbidity indices.

**Figure S5.** Calibration curves for three-year mortality across comorbidity indices

**Figure S6.** Calibration curves for five-year mortality across comorbidity indices

**Figure S7.** Calibration performance across demographic and socioeconomic subgroups

**Figure S8.** Range of subgroup discrimination across demographic and socioeconomic subgroups

**Figure S9.** Subgroup discrimination (AUROC) across demographic and socioeconomic groups

**Figure S10.** Subgroup-specific condition weights for one-year PCS variants

**Figure S11.** Sensitivity analysis of subgroup-specific condition weights with negative coefficients retained one-year PCS variants.

**Table S1.** Variable definitions

| **Variable** | **Source** | **Definition** | **Coding / Categories** |
| --- | --- | --- | --- |
| Age at admission | Epic Cosmos | Patient age (years) at the index hospital admission | Continuous; Age groups for subgroup models: 18 to 44, 45 to 64, 65 to 79, and ≥80 years |
| Sex | Epic Cosmos | Sex recorded in the EHR | Female, Male |
| Race | Epic Cosmos | Race recorded in the EHR | White, Black or African American, Asian, American Indian or Alaska Native, Native Hawaiian or Other Pacific Islander, Other/Unknown. Used for evaluation only; not included in PCS. |
| Ethnicity | Epic Cosmos | Ethnicity recorded in the EHR | Hispanic or Latino, Non-Hispanic or Latino, Other/Unknown. Used for evaluation only; not included in PCS. |
| Insurance (Financial Class) | Epic Cosmos | Primary payer at index admission | Medicare, Medicaid, Self-pay, Other/Miscellaneous, Unknown |
| Social Vulnerability Index (SVI) | Epic Cosmos (CDC SVI  linkage) | Neighborhood-level social vulnerability measure based on the patient's residential location | Continuous score (0 to 1) |
| Rural-Urban Commuting Area (RUCA) | Epic Cosmos | Geographic classification of patient residence | Original RUCA codes (1-10) were recategorized into four geographic categories (Metropolitan, Micropolitan, Small Town, and Rural) according to the RUCA classification. |
| Comorbid conditions | ICD-10-CM diagnoses mapped to AHRQ CCSR | Chronic comorbid conditions identified from diagnosis codes before the index admission | Binary indicators for clinician-selected chronic CCSR categories |
| One-year mortality | Epic Cosmos | Death occurring within 365 days of index admission | Binary outcome |
| Three-year mortality | Epic Cosmos | Death occurring within 1095 days of index admission | Binary outcome |
| Five-year mortality | Epic Cosmos | Death occurring within 1825 days of index admission | Binary outcome |

Abbreviations: CCSR, Clinical Classifications Software Refined; EHR, electronic health record; ICD-10-CM, International Classification of Diseases, Tenth Revision, Clinical Modification; PCS, Personalized Comorbidity Score; RUCA, Rural-Urban Commuting Area; SVI, Social Vulnerability Index.

**Table S2.** Model discrimination for 1-year mortality

| **Model** | **AUROC (95% CI)** | **C-statistic (95% CI)** |
| --- | --- | --- |
| Original CCI | 0.714 (0.707-0.721) | 0.707 (0.700-0.713) |
| Quan CCI | 0.719 (0.712-0.726) | 0.712 (0.705-0.719) |
| VW ECI | 0.730 (0.723-0.737) | 0.722 (0.716-0.729) |
| PCS Core (sensitivity analysis) | 0.822 (0.816-0.827); p<0.001 | 0.812(0.806-0.817); p<0.001 |
| PCS Core | 0.812 (0.806-0.817); p<0.001 | 0.802 (0.796-0.807); p<0.001 |
| PCS Extended (sensitivity analysis) | 0.822 (0.817-0.828); p<0.001 | 0.812 (0.807-0.817); p<0.001 |
| PCS Extended | 0.813 (0.807-0.818); p<0.001 | 0.803 (0.797-0.808); p<0.001 |

Abbreviations: AUROC, area under the receiver operating characteristic curve; CI, confidence interval; CCI, Charlson Comorbidity Index; ECI, Elixhauser Comorbidity Index; PCS, Personalized Comorbidity Score. AUROC and C-statistic were evaluated in the held-out SneakPeek test cohort. Confidence intervals and *P* values were derived from 2000 bootstrap iterations. Original CCI served as the reference comparator. Sensitivity analyses retained negative regression coefficients rather than constraining them to zero.

**Table S3.** Model discrimination for 3-year mortality

| **Model** | **AUROC (95% CI)** | **C-statistic (95% CI)** |
| --- | --- | --- |
| Original CCI | 0.709 (0.704-0.715) | 0.697 (0.692-0.702) |
| Quan CCI | 0.707 (0.702-0.712) | 0.695 (0.691-0.701) |
| VW ECI | 0.715 (0.710-0.721) | 0.704 (0.698-0.709) |
| PCS Core (sensitivity analysis) | 0.808 (0.804-0.813); p<0.001 | 0.791 (0.787-0.795); p<0.001 |
| PCS Core | 0.799 (0.795-0.804); p<0.001 | 0.782 (0.778-0.787); p<0.001 |
| PCS Extended (sensitivity analysis) | 0.807 (0.802-0.811); p<0.001 | 0.790 (0.786-0.794); p<0.001 |
| PCS Extended | 0.800 (0.795-0.804); p<0.001 | 0.783 (0.779-0.787); p<0.001 |

Abbreviations: AUROC, area under the receiver operating characteristic curve; CI, confidence interval; CCI, Charlson Comorbidity Index; ECI, Elixhauser Comorbidity Index; PCS, Personalized Comorbidity Score. AUROC and C-statistic were evaluated in the held-out SneakPeek test cohort. Confidence intervals and *P* values were derived from 2000 bootstrap iterations. Original CCI served as the reference comparator. Sensitivity analyses retained negative regression coefficients rather than constraining them to zero.

**Table S4.** Model discrimination for 5-year mortality

| **Model** | **AUROC (95% CI)** | **C-statistic (95% CI)** |
| --- | --- | --- |
| Original CCI | 0.706 (0.701-0.711) | 0.690 (0.685-0.694) |
| Quan CCI | 0.695 (0.690-0.699) | 0.681 (0.677-0.686) |
| VW ECI | 0.707 (0.703-0.712) | 0.692 (0.688-0.697) |
| PCS Core (sensitivity analysis) | 0.802 (0.798-0.806);p<0.001 | 0.779 (0.776-0.783); p<0.001 |
| PCS Core | 0.795 (0.790-0.799); p<0.001 | 0.772 (0.768-0.775); p<0.001 |
| PCS Extended (sensitivity analysis) | 0.801 (0.797-0.805); p<0.001 | 0.778 (0.774-0.782); p<0.001 |
| PCS Extended | 0.796 (0.792-0.800); p<0.001 | 0.773 (0.769-0.777); p<0.001 |

Abbreviations: AUROC, area under the receiver operating characteristic curve; CI, confidence interval; CCI, Charlson Comorbidity Index; ECI, Elixhauser Comorbidity Index; PCS, Personalized Comorbidity Score. AUROC and C-statistic were evaluated in the held-out SneakPeek test cohort. Confidence intervals and *P* values were derived from 2000 bootstrap iterations. Original CCI served as the reference comparator. Sensitivity analyses retained negative regression coefficients rather than constraining them to zero.

**Table S5.** Characteristics of the Stanford Health Care validation cohort

|  | **Missing** | **Overall (n=82,947)** |
| --- | --- | --- |
| Age at admission, mean (SD) | 0 | 60.6 (17.9) |
| Age group, n (%) | 0 |  |
| 18-44 |  | 16,728 (20.2) |
| 45-64 |  | 28,810 (34.7) |
| 65-79 |  | 26,046 (31.4) |
| 80+ |  | 11,363 (13.7) |
| Sex, n (%) | 0 |  |
| Female |  | 41,842 (50.4) |
| Male |  | 41,105 (49.6) |
| Insurance, n (%) | 14,505 |  |
| Medicaid |  | 15,414 (22.5) |
| Medicare |  | 12,931 (18.9) |
| Miscellaneous/Other |  | 40,097 (58.5) |
| Race, n (%) | 19 |  |
| Asian |  | 11,588 (14.0) |
| Black |  | 3798 (4.6) |
| Patient refused |  | 658 (0.8) |
| Hawaiian or Pacific Islander |  | 1040 (1.3) |
| Native American |  | 372 (0.4) |
| Other |  | 15697 (18.9) |
| Unknown |  | 922 (1.1) |
| White |  | 48,853 (58.9) |
| Ethnicity, n (%) | 14 |  |
| Declines to State |  | 717 (0.9) |
| Hispanic/Latino |  | 12,982 (15.7) |
| Non-Hispanic/Non-Latino |  | 68,391 (82.5) |
| Unknown |  | 844 (1.0) |
| Social Vulnerability Index, mean (SD) |  |  |
| Overall | 197 | 0.63 (0.11) |
| Household Characteristics | 197 | 0.54 (0.14) |
| Housing/Transportation | 197 | 0.73 (0.11) |
| Socioeconomic | 197 | 0.41 (0.24) |
| Racial Ethnic Minority Status | 197 | 0.83 (0.09) |
| Rural Urban Category | 167 |  |
| Metropolitan |  | 74,734 (90.1%) |
| Micropolitan |  | 60 (0.1%) |
| Small Town/Rural |  | 7986 (9.6%) |
| One year mortality, n (%) | 0 |  |
| 1 |  | 9595 (11.6) |

Because Stanford Health Care provides residential ZIP codes at the three-digit level only, neighborhood-level measures were approximated at the three-digit ZIP code level. Mean Social Vulnerability Index (SVI) values and the modal Rural-Urban Commuting Area (RUCA) category were assigned to each three-digit ZIP code. Cosmos financial class was obtained directly from the index hospital admission record. Stanford financial class was derived by linking inpatient admissions to coverage data using coverage effective dates at admission; admissions without a matched coverage record (~17%) were classified as missing prior to analysis and recoded to Miscellaneous/Other for modeling (median value).

**Table S6.** Characteristics of the MIMIC-IV validation cohort

|  | **Missing** | **Overall (n=108,922)** |
| --- | --- | --- |
| Age at admission, mean (SD) | 0 | 60.91 ± 18.75 |
| Age group, n (%) | 0 |  |
| 18-44 |  | 35,239 (32.4%) |
| 45-64 |  | 22,143 (20.3%) |
| 65-79 |  | 33,056 (30.3%) |
| 80+ |  | 18,484 (17.0%) |
| Insurance, n (%) | 2110 |  |
| Medicaid |  | 18,847 (17.3%) |
| Medicare |  | 48,612 (44.6%) |
| Miscellaneous /Other |  | 39,353 (36.2%) |
| Race/Ethnicity, n (%) | 0 |  |
| White |  | 71,391 (65.5%) |
| Black |  | 14,168 (13.0%) |
| Unknown |  | 8,562 (7.9%) |
| Hispanic/Latino |  | 5,274 (4.8%) |
| Other |  | 4,605 (4.2%) |
| Asian |  | 4,520 (4.1%) |
| Native American |  | 255 (0.2%) |
| Hawaiian or Pacific Islander |  | 147 (0.1%) |
| Sex, n (%) | 0 |  |
| Male |  | 54,791 (50.3%) |
| Female |  | 54,131 (49.7%) |
| 1-year mortality, n (%) | 0 |  |
| 1 |  | 13,260 (12.2%) |

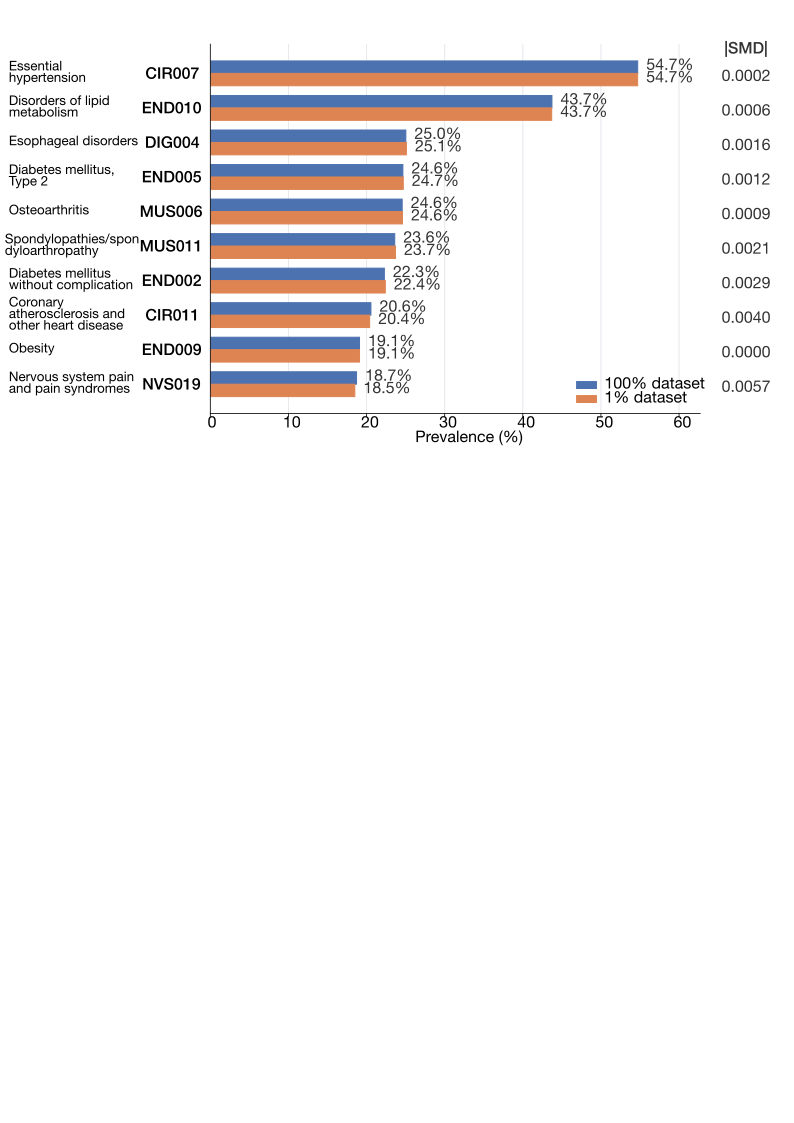

**Figure S1.** Ten most prevalent CCSR-defined chronic comorbidities in the Epic Cosmos cohort

The prevalence of the ten most common clinician-selected chronic conditions is shown for the full development cohort (100% dataset) and the held-out SneakPeek test cohort (1% dataset). Standardized mean differences (SMDs) are presented to assess the representativeness of the SneakPeek subset relative to the full cohort.

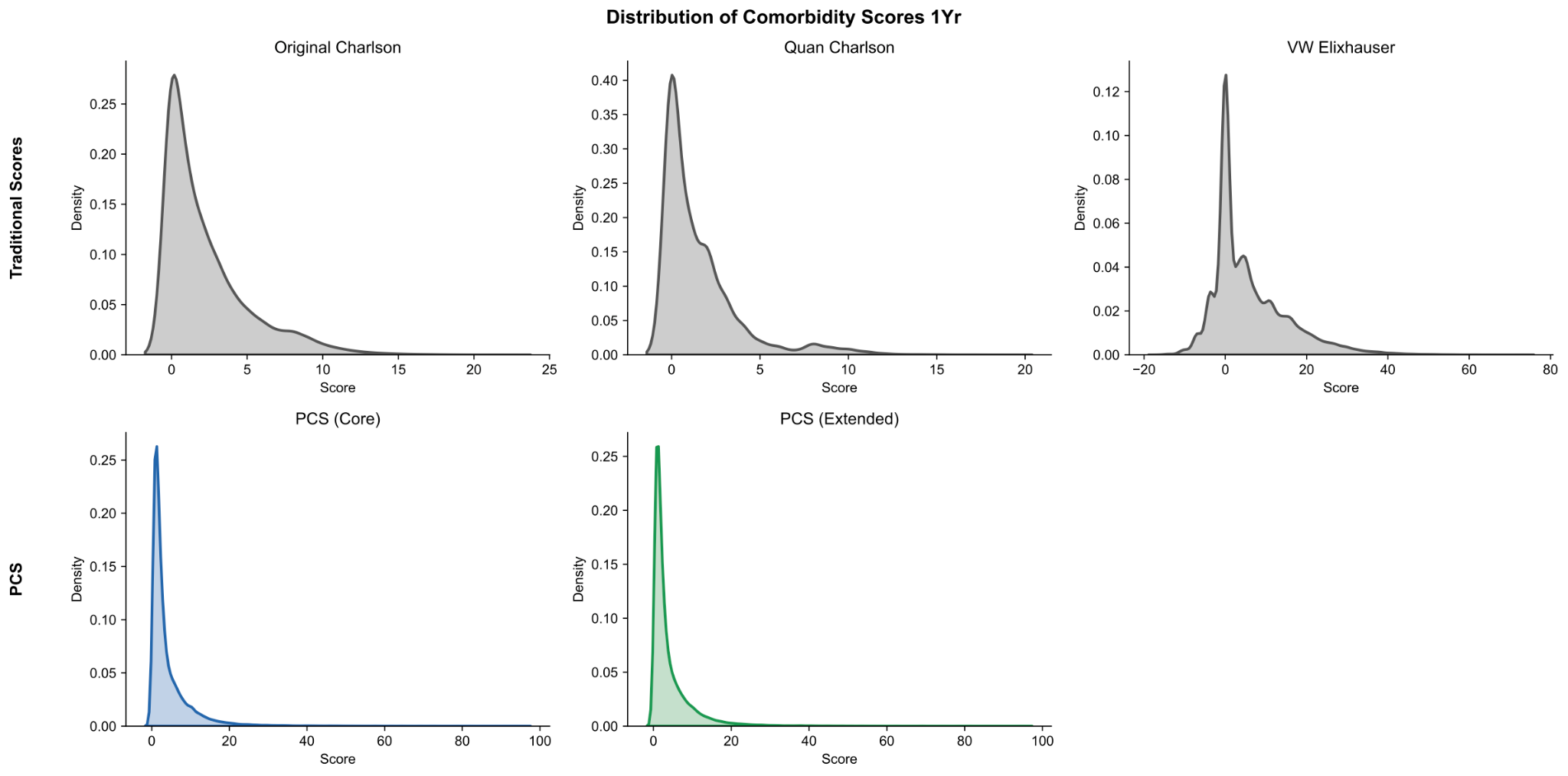

**Figure S2** Score distributions for traditional comorbidity indices and one-year PCS variants in the SneakPeek 1% held-out validation dataset.

Kernel density plots illustrate the distribution of one-year comorbidity scores for the Original Charlson Comorbidity Index, Quan Charlson Comorbidity Index, van Walraven Elixhauser Comorbidity Index, PCS Core, and PCS Extended in the held-out SneakPeek test cohort.

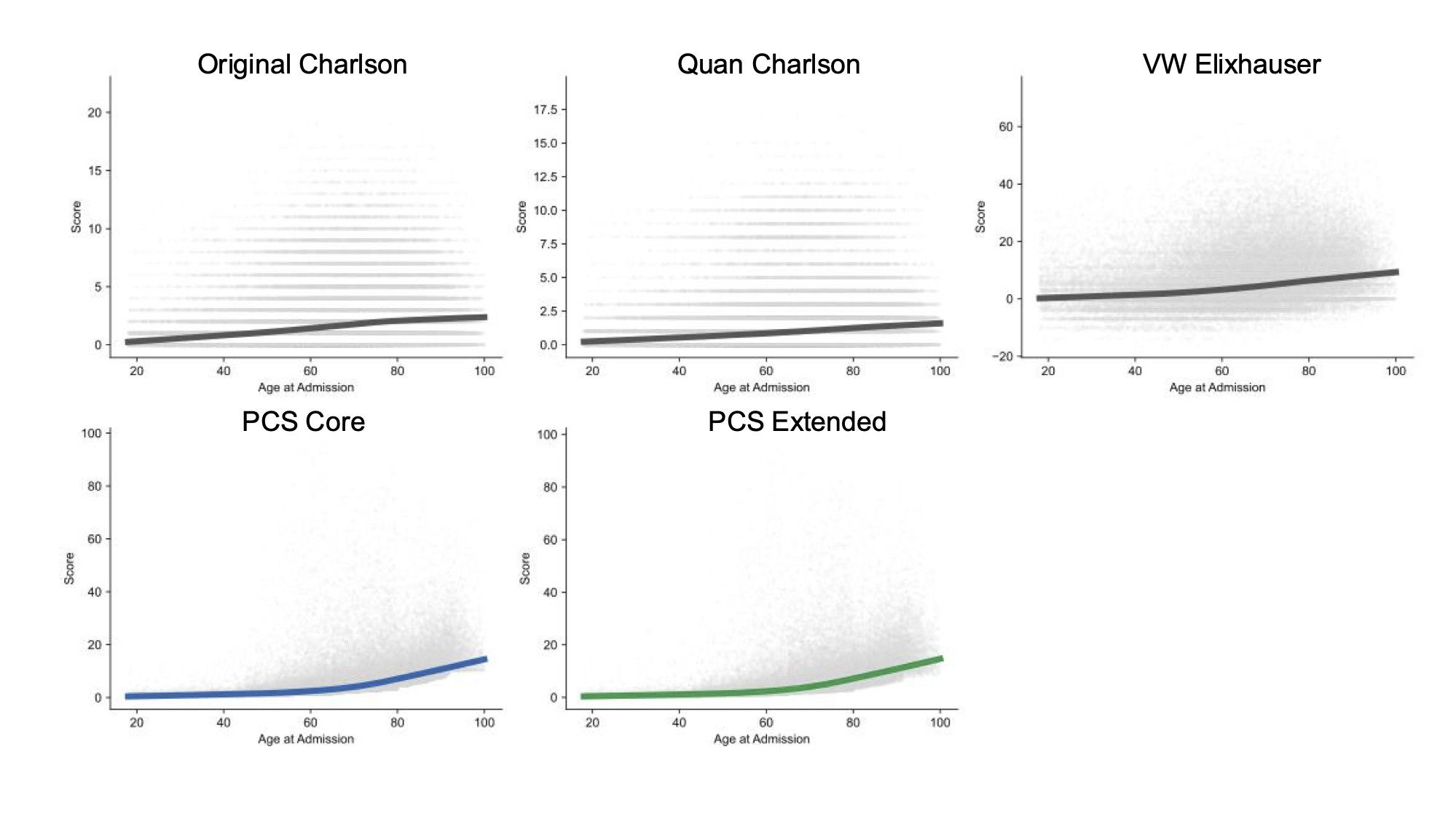

**Figure S3.** Relationship between age at admission and comorbidity scores across comorbidity indices.

Scatterplots show individual comorbidity scores by age at admission in the held-out SneakPeek test cohort. Solid lines represent locally weighted scatterplot smoothing (LOWESS) trends. Traditional indices (Original Charlson Comorbidity Index, Quan Charlson Comorbidity Index, and van Walraven Elixhauser Comorbidity Index) are compared with PCS Core and PCS Extended.

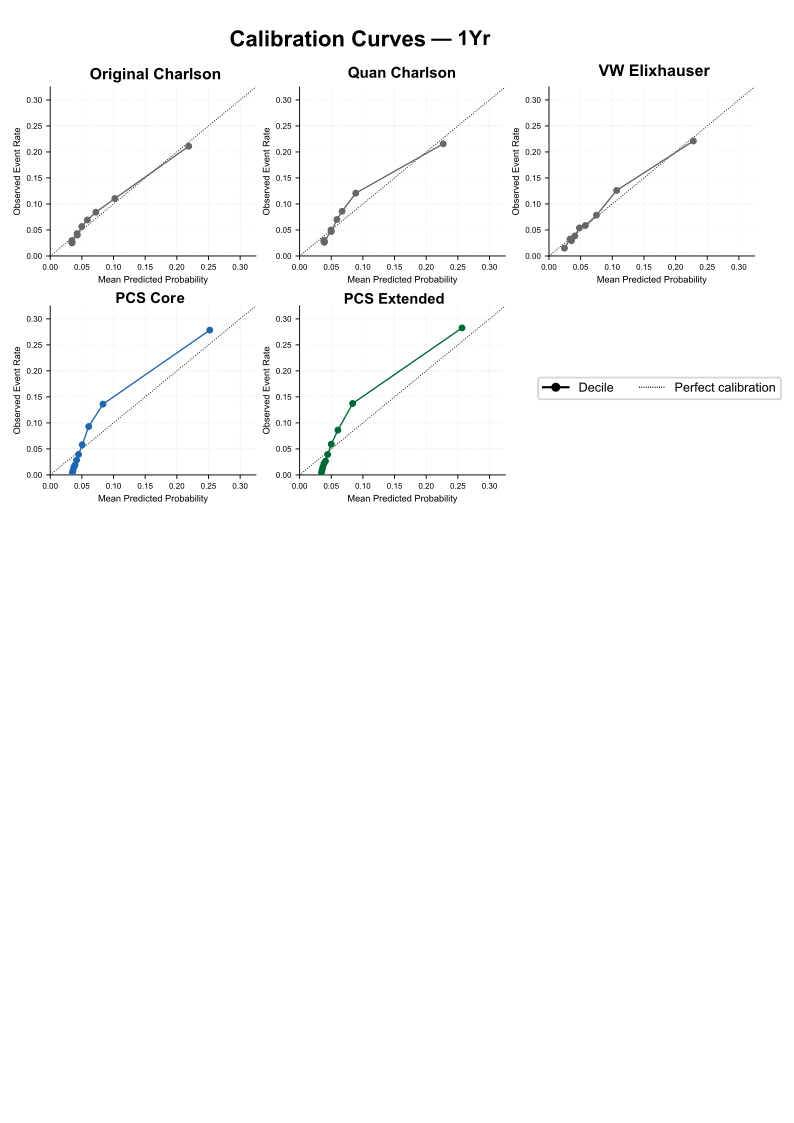

**Figure S4. Calibration curves for one-year mortality across comorbidity indices.**

Calibration was assessed using deciles of predicted risk in the held-out SneakPeek test cohort. Points represent observed versus predicted event rates within each decile after Platt scaling, with the diagonal dashed line indicating perfect calibration. Traditional comorbidity indices (Original Charlson Comorbidity Index, Quan Charlson Comorbidity Index, and van Walraven Elixhauser Comorbidity Index) are compared with PCS Core and PCS Extended.

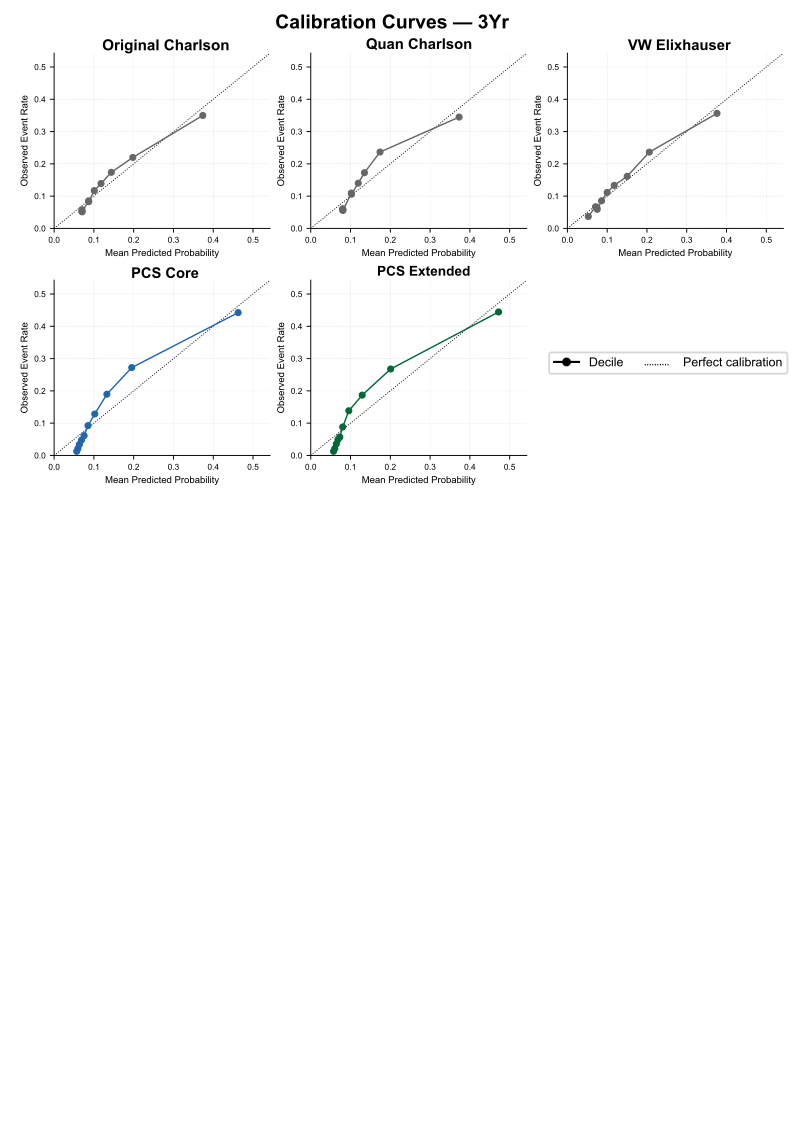

**Figure S5. Calibration curves for three-year mortality across comorbidity indices.**

Calibration was assessed using deciles of predicted risk in the held-out SneakPeek test cohort. Points represent observed versus predicted event rates within each decile after Platt scaling, with the diagonal dashed line indicating perfect calibration. Traditional comorbidity indices (Original Charlson Comorbidity Index, Quan Charlson Comorbidity Index, and van Walraven Elixhauser Comorbidity Index) are compared with PCS Core and PCS Extended.

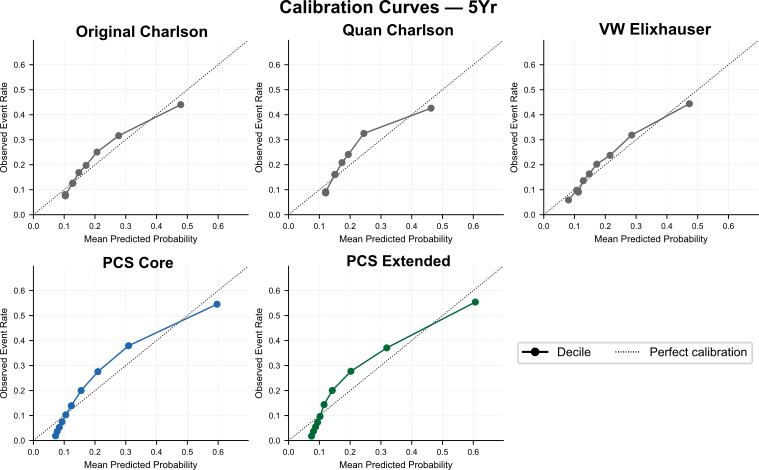

**Figure S6. Calibration curves for five-year mortality across comorbidity indices.**

Calibration was assessed using deciles of predicted risk in the held-out SneakPeek test cohort. Points represent observed versus predicted event rates within each decile after Platt scaling, with the diagonal dashed line indicating perfect calibration. Traditional comorbidity indices (Original Charlson Comorbidity Index, Quan Charlson Comorbidity Index, and van Walraven Elixhauser Comorbidity Index) are compared with PCS Core and PCS Extended.

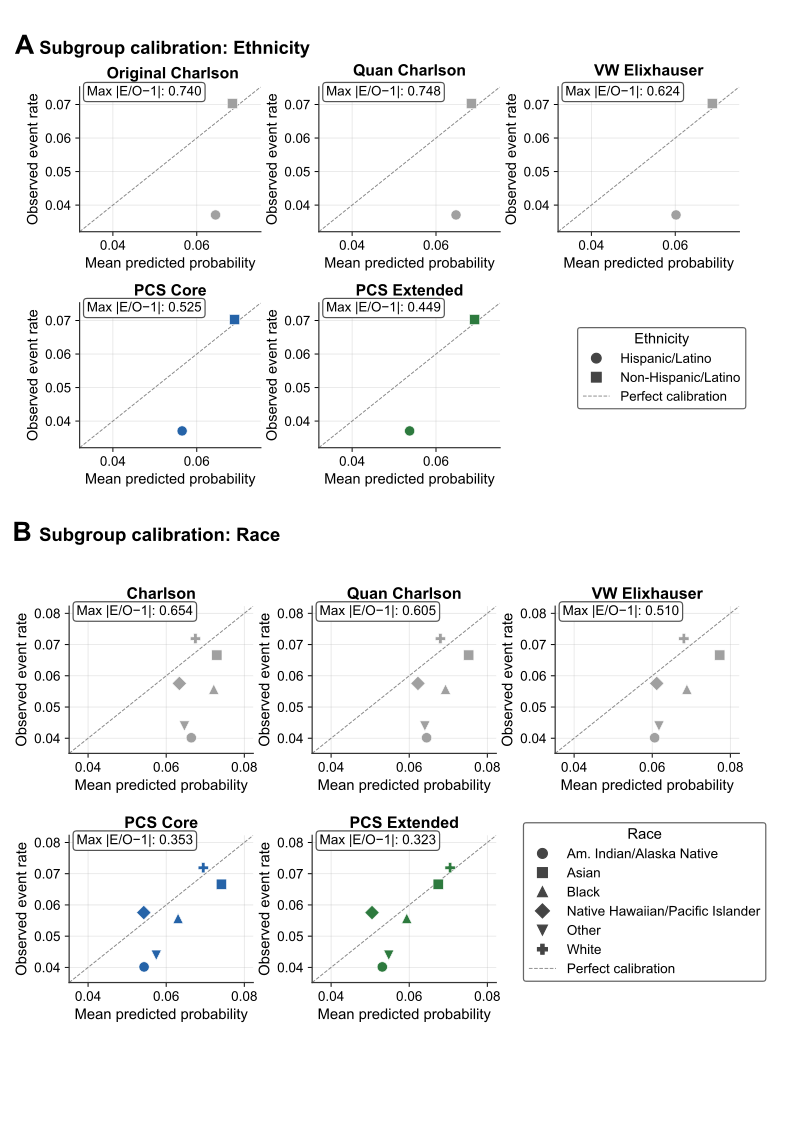

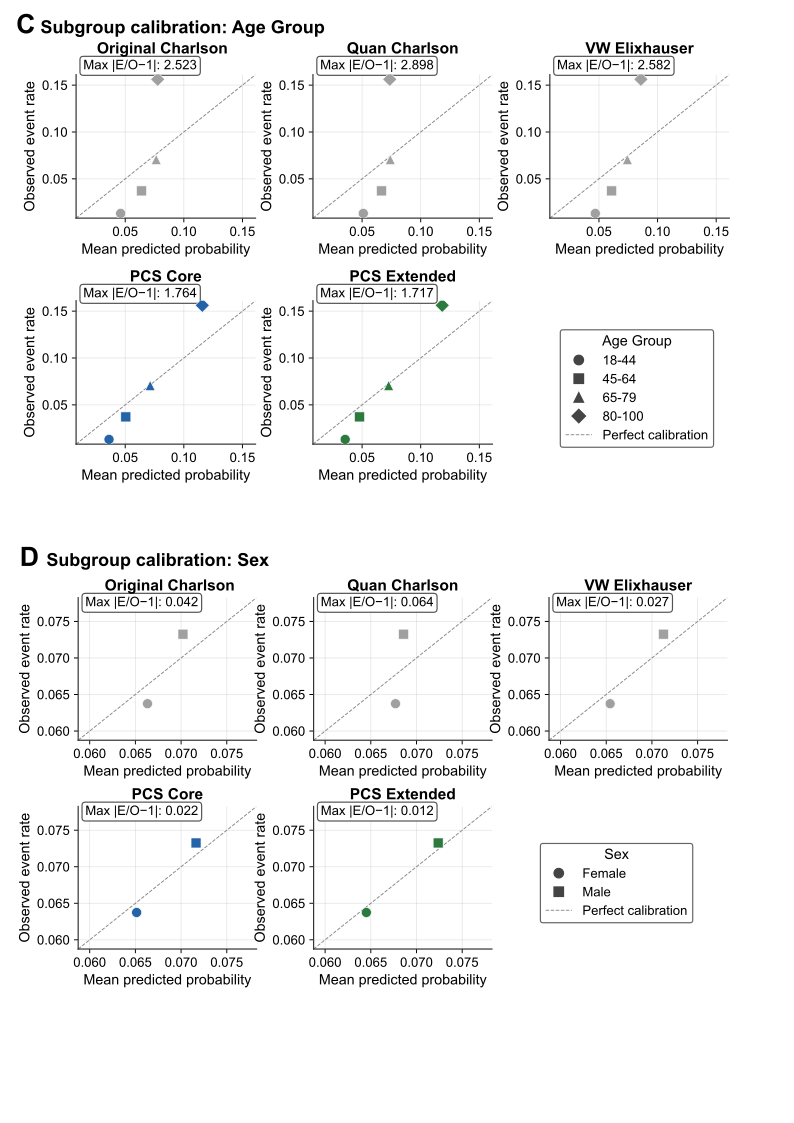

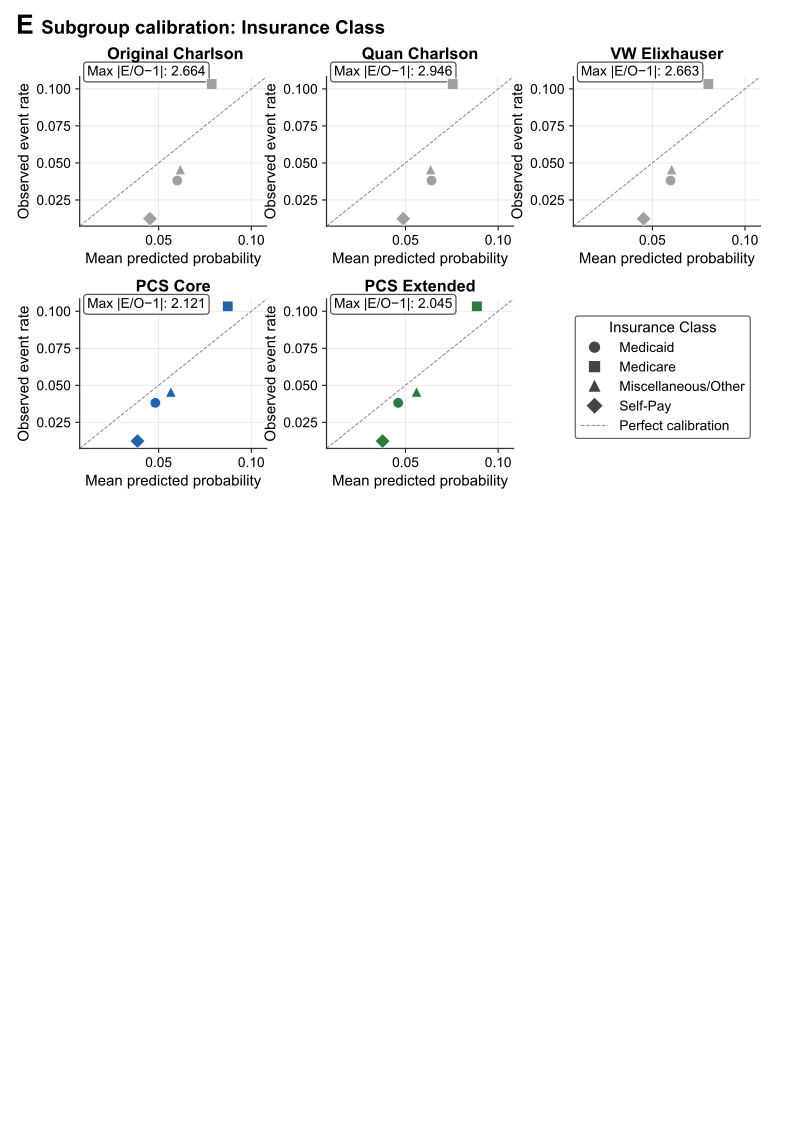

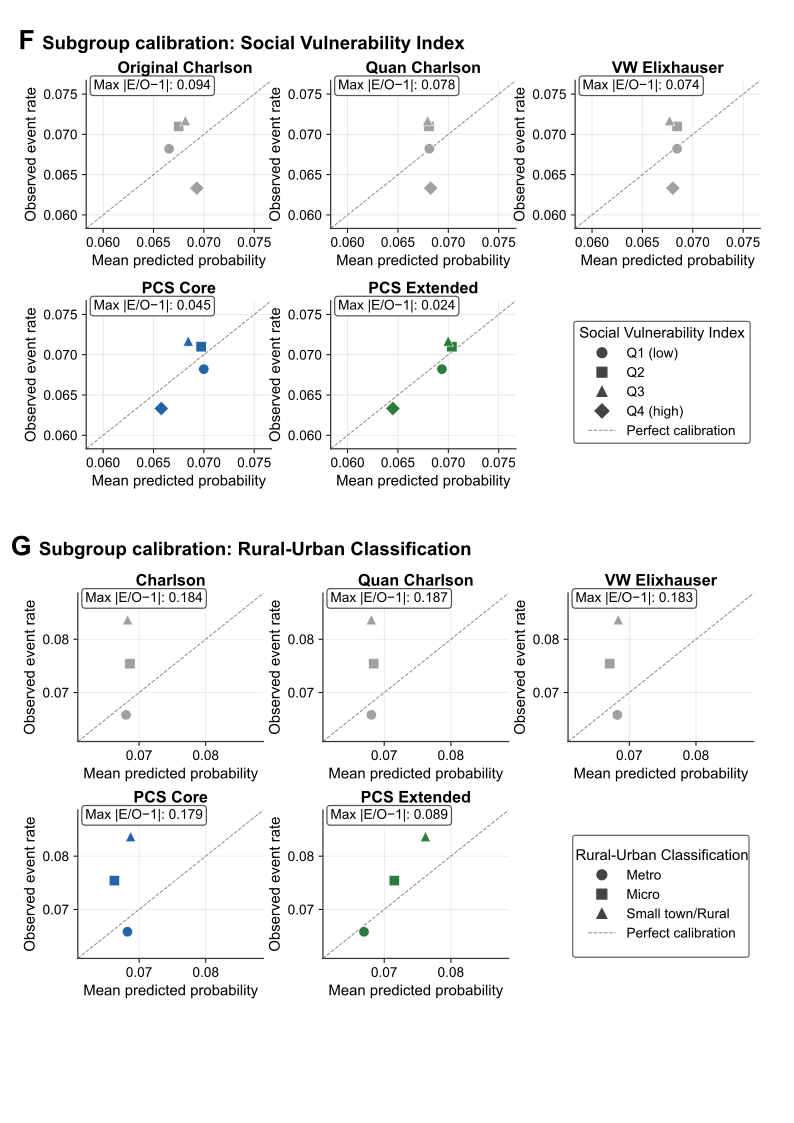

**Figure S7.** Calibration performance across demographic and socioeconomic subgroups**.**

Calibration plots compare the Original Charlson Comorbidity Index, Quan Charlson Comorbidity Index, van Walraven Elixhauser Comorbidity Index, PCS Core, and PCS Extended across demographic and socioeconomic subgroups after Platt scaling. Each point represents the observed event rate versus the mean predicted probability for a subgroup. The dashed diagonal line indicates perfect calibration. The maximum absolute calibration error (Max |E/O − 1|) is shown for each model. (A) Ethnicity. (B) Race. (C) Age group. (D) Sex. (E) Insurance class. (F) Social Vulnerability Index (SVI) quartiles. (G) Rural-Urban Commuting Area (RUCA) classification.

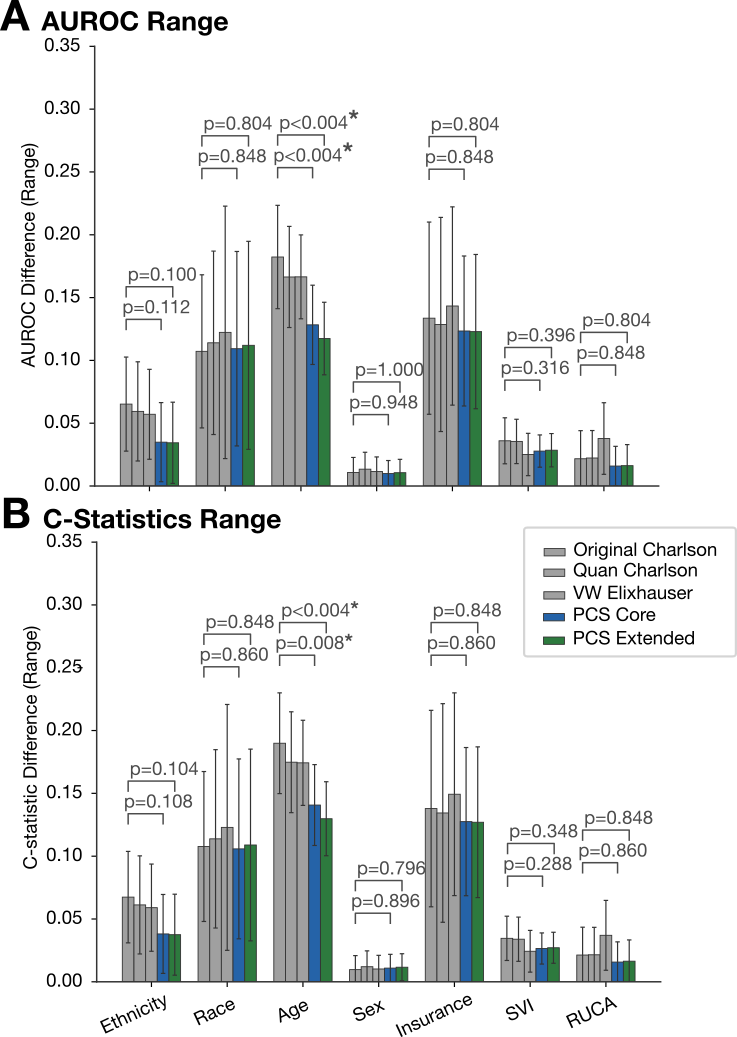

**Figure S8.** Range of subgroup discrimination across demographic and socioeconomic subgroups.

(A) AUROC range and (B) C-statistic range across age, sex, race, ethnicity, insurance class, Social Vulnerability Index (SVI) quartiles, and Rural-Urban Commuting Area (RUCA) classification for the Original Charlson Comorbidity Index, Quan Charlson Comorbidity Index, van Walraven Elixhauser Comorbidity Index, PCS Core, and PCS Extended. Error bars represent 95% confidence intervals derived from 500 bootstrap iterations. P values compare PCS Core and PCS Extended with the best-performing traditional comorbidity index within each subgroup category. Because statistical significance was estimated using 500 bootstrap iterations, the smallest attainable two-sided *P* value is 0.004. *P* < 0.05 was considered statistically significant.

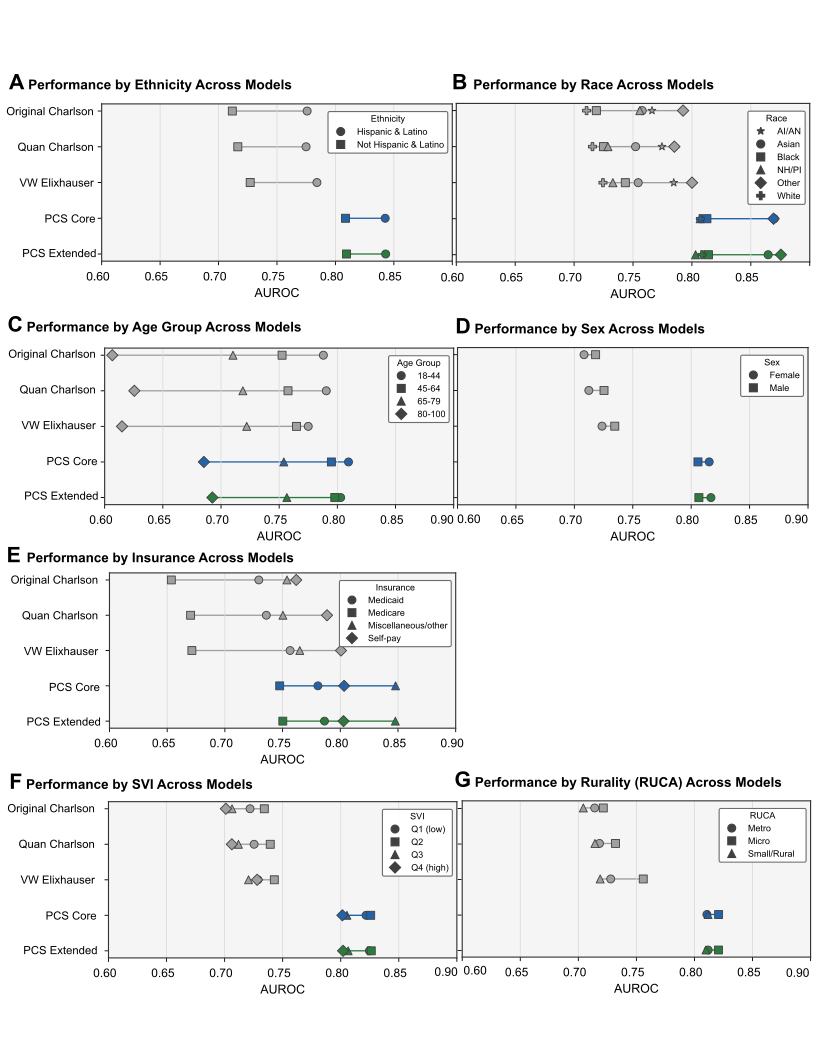

**Figure S9.** Subgroup discrimination (AUROC) across demographic and socioeconomic groups

AUROC for 1-year mortality is shown for Original Charlson, Quan Charlson, VW Elixhauser, PCS Core, and PCS Extended within subgroups defined by (A) ethnicity, (B) race, (C) age group, (D) sex, (E) insurance class, (F) Social Vulnerability Index (SVI) quartile, and (G) Rural-Urban Commuting Area (RUCA) classification. Each point represents the AUROC for a given subgroup, connected by lines within each model to illustrate the spread of discrimination

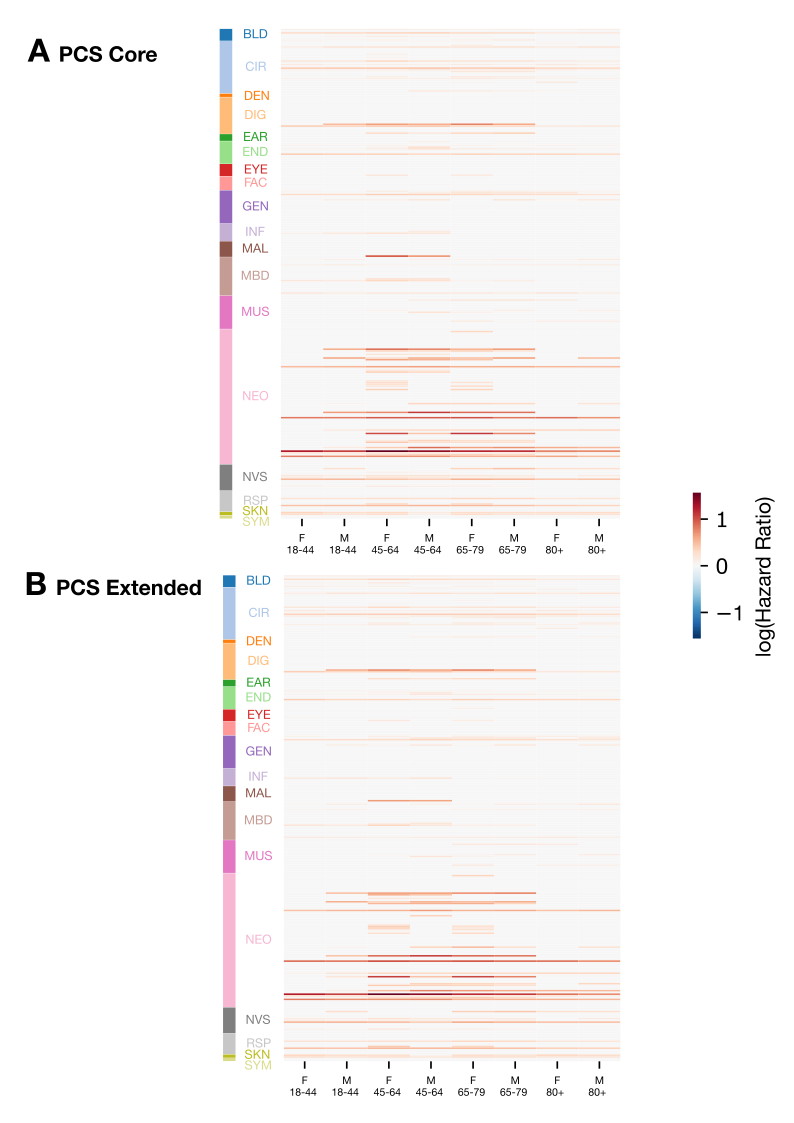

**Figure S10.** Subgroup-specific condition weights for one-year PCS variants

Heatmaps display log(hazard ratio) for each CCSR-defined comorbidity condition (rows) across eight age-sex subgroups (columns), grouped by CCSR body system category (left color bar). Negative coefficients were set to zero in this primary model specification. Darker red indicates a stronger positive association with mortality risk; gray indicates a coefficient of zero or near zero.

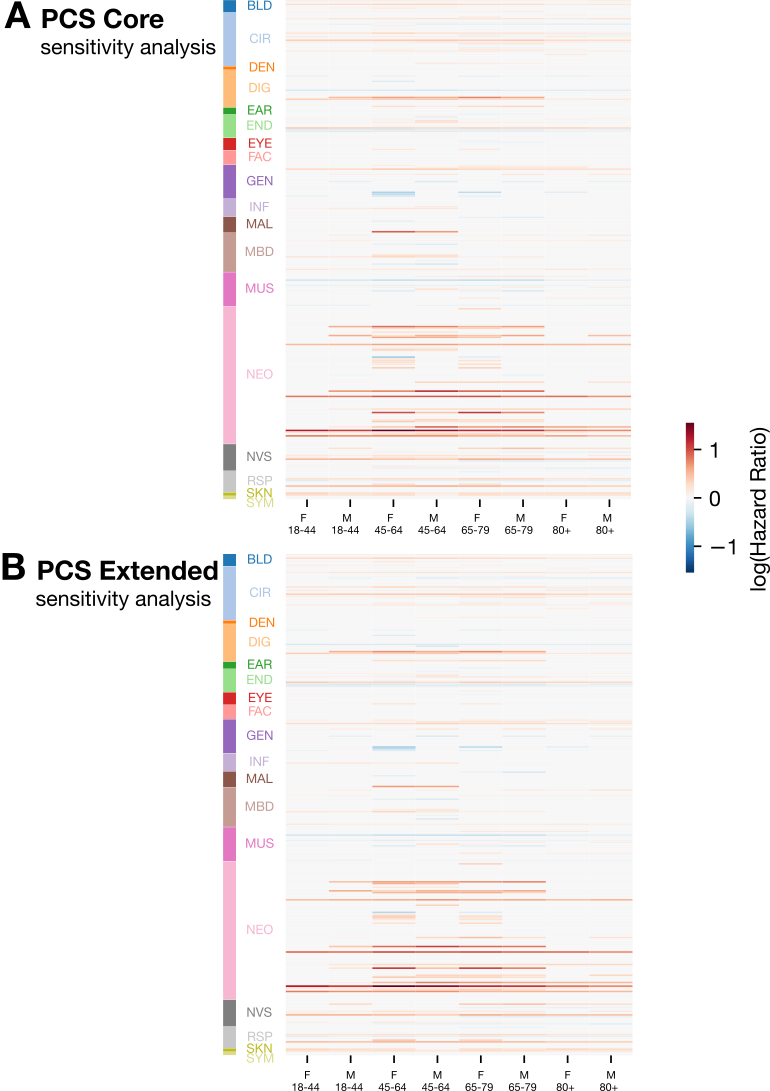

**Figure S11.** Sensitivity analysis of subgroup-specific condition weights with negative coefficients retained one-year PCS variants.

Heatmaps display log(hazard ratio) for each CCSR-defined comorbidity condition (rows) across eight age-sex subgroups (columns), grouped by CCSR body system category (left color bar). Unlike the primary specification, negative coefficients were not constrained to zero. Blue indicates a negative association with mortality risk; red indicates a positive association.
